# Early-life gut microbiome is associated with immune response to the oral rotavirus vaccine in healthy infants in the US

**DOI:** 10.1101/2025.09.22.25336338

**Authors:** Janiret Narváez Miranda, Michael B. Sohn, Daniel Velasquez-Portocarrero, Kelechi Ejiofor, Ann L. Gill, Robert Beblavy, Xing Qiu, Nathan Laniewski, Jessica Brunner, Meghan Best, Alena Leger, Allison Macomber, Sarah L. Caddy, Baoming Jiang, Tom O’Connor, Steven R. Gill, Kristin Scheible

## Abstract

**Background:** Rotavirus remains a leading cause of childhood mortality worldwide, despite the widespread introduction of oral rotavirus vaccines (ORVs). While emerging evidence supports a link between microbiome and vaccine response, findings have been inconsistent, especially across geographic and socioeconomic contexts, and none have been conducted in a US-based cohort. This study investigates the development of the infant gut microbiome and its association with immunogenicity following RotaTeq administration in U.S. infants.

**Methods:** We conducted a longitudinal analysis of infants in Rochester, New York, using 16S rRNA sequencing data to assess microbiome composition. We used rotavirus-specific immunoglobulin A (Rotavirus-IgA) titers at the sixth-month study visit (M6) in plasma to determine the seroresponse to vaccination. Clinical metadata were used to evaluate the influence of different factors on microbial diversity over the first year of life and Rotavirus-IgA titers at the M6 visit. Microbiome data from the M1 visit and Rotavirus-IgA at the M6 visit were used to assess the relationship between the infant gut microbiome and ORV immune responses.

**Findings:** The infant gut microbiome followed characteristic developmental patterns during the first year (N=264). At the M6 visit, 65 infants had a Rotavirus-IgA geometric mean titer of 455, 95% CI:[272-761]. In a sub-cohort that included the complete dataset of immunogenicity and microbiome (N=47), higher alpha diversity at the month 1 (M1) visit was significantly associated with higher Rotavirus-IgA titers at the M6 visit (ß= 2.151, 95% CI:[0.31-3.99], p=0.023). Specific taxa present at the M1 visit, including *Collinsella* (ß: 0.243, 95% CI:[0.076, 0.392], q= 0.037), *Atopobium* (ß: 0.262, 95% CI:[0.066, 0.458], q= 0.062), and *Schaalia radingae* (ß: 0.28, 95% CI:[0.116, 0.458], q=0.018), were positively associated with Rotavirus-IgA titers. In contrast, *Bifidobacterium* (ß: −0.204,95% CI:[−0.323, −0.085], q=0.012), *Lactobacillus* (ß: −0.17, 95% CI:[− 0.314, −0.035], q= 0.087), *Klebsiella* (ß: −0.195, 95% CI:[−0.331, −0.058], q= 0.042), *Escherichia-Shigella* (ß: - 0.128, 95% CI:[−0.245, −0.012], q= 0.162), *Streptococcus salivarius* (ß: −0.229, 95% CI:[−0.359, −0.098], q= 0.012), and *Peptostreptococcus anaerobius* (ß: −0.176, 95% CI:[−0.338, −0.014], q= 0.162) were negatively associated.

**Interpretation:** In a healthy U.S.-infant cohort, we report a significant association between the early-life infant gut microbiome and RotaTeq-vaccinated infants’ Rotavirus-IgA titers. This study contributes to a clearer understanding of microbiome-vaccine interactions, particularly in high-income settings where existing evidence has been limited.

**Funding:** Office of the Director of the National Institutes of Health, National Institute of Mental Health of the National Institutes of Health, and the National Center for Advancing Translational Sciences of the National Institutes of Health

## Introduction

Diarrheal diseases remain a leading cause of child mortality worldwide, and Rotavirus is a major contributor across all age groups.^1^ Global introduction of the oral live-attenuated rotavirus vaccine (ORVs) to infants 2-8 months of age resulted in a drop in severe pediatric gastroenteritis and child death cases.^2^ ORV’s immunogenicity exhibits geographical variability^3^, with lower responses reported in low-middle-income countries (LMICs) compared to high-income countries (HICs).

Several factors can influence ORV immune responses, including Rotavirus prevalence, transfer of maternal antibodies, and co-administration of other oral vaccines.^4–6^ For example, maternal antibodies transferred through breastmilk have been shown to exert an influence on ORV immune responses in high Rotavirus-prevalence LMIC.^7^ In contrast, in the US, no effect of breastmilk antibodies on ORV responses has been observed.^8^ Another factor hypothesized to influence ORV immune responses is the gut microbiome. The gut environment, where ORV replication stimulates immunity against natural infection, hosts a microbiome that is essential for immune development, and dysbiosis predicts adverse immune-related outcomes such as allergies and diabetes.^9,10^ While the microbiota-immune systems interaction is incompletely understood, bacterial members of the microbiome produce metabolites (e.g., short-chain fatty acids)^11^ and antigens (e.g., polysaccharide A)^12^ that interact with innate and adaptive immune cells, ultimately influencing IgA production in the gut.

Several studies have aimed to identify an association between the developing infant gut microbiome and antigen-specific responses, such as to ORV. Most studies have been conducted in LMICs, where the ORV immunogenicity is markedly variable and confounded by high Rotavirus prevalence. Only three studies have included HIC as comparison groups, and the single study among those three that characterized both ORV immune responses and gut microbiome had null findings.^14,16,17^ Heterogeneity between study populations, and limitations posed by the compositional nature of microbiome data, have led to inconclusive results regarding the independent effect of gut microbiome on ORV immunogenicity.^13–20^ We hypothesized that by leveraging a US population with high vaccine uptake, low Rotavirus infection prevalence, prospectively collected breastfeeding behavior, we could better isolate the effect of infant gut microbiome on ORV immune responses. In this study, we employ a rigorous longitudinal approach using analytic methods that address the limitation of relative abundance measures, to characterize microbiome composition, vaccine responses, and their relationship in early life.

## Methods

### Study description

From December 2015 to April 2019, pregnant participants were recruited from outpatient obstetrics clinics affiliated with the University of Rochester Medical Center (URMC). Pregnant subjects and their newborns were enrolled as part of the Understanding Pregnancy Signals and Infant Development and Environmental Influences on Child Health Outcomes (UPSIDE-ECHO) study.^21^ The study protocol was approved by the University of Rochester School of Medicine and Dentistry Institutional Review Board (#58456), and all participants provided written informed consent. Eligibility criteria for pregnant participants included: <14 weeks of gestation, age 18 or older, singleton pregnancy, no known substance problems or a history of psychotic illness, and ability to communicate in English. Exclusion criteria included women with major endocrine disorders (e.g., polycystic ovary syndrome, pre-pregnancy diabetes), high-risk pregnancies, or significant obstetric problems. Infants born before 37 weeks of gestation were excluded from the postnatal study phases.

Study visits occurred in private clinic rooms and included biospecimen collection, and questionnaires. Maternal (e.g., race/ethnicity, education) and infant characteristics (e.g., sex, race/ethnicity, antibiotic use, diet) were obtained from maternal self-reports and medical records. Postnatal follow-up occurred at one (M1), six (M6), and twelve (M12) months. Infant rectal swabs were collected at birth, M1, M6, and M12 for microbiome sequencing. Blood samples for plasma were collected in EDTA at birth (cord blood) and at M6 and M12.

Rotavirus vaccination followed CDC guidelines and standard URMC practice. Vaccines were administered orally: RotaTeq (three doses at two, four, six months) or Rotarix (two doses at two and four months). The first dose was recommended before 15 weeks, with full vaccination completed before eight months of age.

### DNA Extraction and 16S rRNA amplicon sequencing

Total genomic DNA was extracted from rectal samples by modifying the Zymo Quick-DNA™ Fecal/Soil Microbe Miniprep Kit. The V3–V4 regions of the 16S rRNA gene were amplified using dual-indexed primers and pooled libraries were sequenced (2□×□312 bp) on an Illumina MiSeq platform at the University of Rochester Genomics Research Center. Each sequencing run included positive, negative, and extraction controls **(appendix p 3)**. Reads were demultiplexed and processed in QIIME2 v2022.2 using DADA2 for denoising and merging. Forward and reverse reads were truncated at 275 bp and 260 bp, respectively, and quality filtering parameters are detailed in the (**appendix p 3)**. Taxonomic classification was performed with a custom Bayesian classifier trained on the SILVA database. Microbiome data were further processed to reduce noise and improve the reliability of any subsequent analysis **(appendix p 4)**

### Rotavirus Vaccine Immune-Outcomes

An initial data pull from the New York State vaccine records indicated that 273 of 306 (89%) infants enrolled in the UPSIDE-ECHO study had received at least one dose of an ORV during their first year of life. The primary immunological outcome of this study was plasma Rotavirus-IgA titers, measured through a RotaTeq antigen-specific Enzyme-Linked Immunoassay (EIA)^22^ (**appendix p 5**).

The coprimary outcome was Rotavirus-IgA seroresponse to RotaTeq or Rotarix, assessed by seroconversion and Rotavirus-IgA titers, expressed as log2-transformed values and geometric mean titers (GMTs). Rotavirus-IgA seropositivity was defined as a titer ≥40. Seroconversion was defined as either (a) seronegative at baseline (titer <40) and seropositive at follow-up, or (b) seropositive at baseline with a ≥4-fold rise in titer at follow-up. Because only 3 of 36 (8%) infants in the immunogenicity–microbiome subcohort were seropositive at birth, baseline antibody levels were considered very low. Seroresponse was therefore evaluated at M6 visit using both log2-transformed titers and GMTs.

### Statistical Analysis

Descriptive statistics were used to summarize population characteristics. Continuous variables were reported as means with standard deviations, and categorical variables as frequencies and percentages. Between-group comparisons used non-parametric or χ^2^ tests as appropriate. Statistical significance was set at p < 0.05, with multiple testing adjusted using the Benjamini–Hochberg procedure. For gut microbiome analyses, alpha diversity was quantified using Shannon’s index and compared across visits and covariates. Longitudinal diversity was modeled using linear mixed-effects models to account for repeated measures, and estimated marginal means were used for pairwise contrasts. Beta diversity was calculated using Bray–Curtis dissimilarity, and overall compositional differences by visit were assessed with permutational multivariate analysis of variance (PERMANOVA). To examine associations between covariates and microbiome diversity, both non-parametric tests (**Supplemental Table 2; appendix p 8**) and regression models adjusting for age and feeding type were applied.

Rotavirus-IgA responses were analyzed longitudinally using linear mixed-effects models with gestational age included as a covariate. Differences in Rotavirus-IgA titer distributions between the full cohort (n = 65) and the sub-cohort (n = 47) across birth, M6, and M12 were evaluated using the Kolmogorov-Smirnov test, while seropositivity and seroconversion rates were compared with Fisher’s exact test. The association between vaccination status at M6 and Rotavirus-IgA titers was assessed using linear regression. A linear regression model was utilized to test the association between vaccination status at the M6 visit and Rotavirus-IgA titers.

Additional multivariable analyses explored relationships between IgA titers and demographic or feeding variables, with effect estimates adjusted for post-conception age.

To investigate links between early microbiome features and vaccine immune outcomes, we modeled the association between microbial alpha diversity at M1 and Rotavirus-IgA titers at M6, adjusting for post-conception age and feeding type. The relationship between overall microbial community composition at M1 and antibody responses was tested using kernel-based association methods. Taxa-level associations with antibody titers were evaluated using penalized regression models designed for compositional microbiome data, and multiple testing correction was applied.

All analyses were performed using RStudio (version 4.5.0). Detailed statistical specifications and implementation steps are provided in the supplementary methods (**appendix p 6-7**).

## Results

### Study Population

From the larger UPSIDE-ECHO cohort, 84.15% of received the ORV vaccine RotaTeq, 12.32% received Rotarix, 2.12% received a mixed vaccine schedule of both RotaTeq and Rotarix, and 1.40% who received none (**Figure 1**). For this study, 177 unique participants were selected from the larger UPSIDE-ECHO cohort based on the availability of plasma samples and vaccination with RotaTeq (**Figure 1**). At birth (n = 147), 54% of infants were female, 57% were identified as White, 75% were vaginally delivered, the mean gestational age was 39.55 ± 1.17 weeks, and the mean weight-for-age z-score was 0.06 ± 0.79 (**Table 1**).

**Table 1.** Study Demographics.

| **Variable** | **Birth**<br>N = 147<sup>1</sup> | **Month Six (M6)**<br>N = 65<sup>1</sup> | **Month Twelve (M12)**<br>N = 60<sup>1</sup> |
| --- | --- | --- | --- |
| **Infant Sex** |  |  |  |
| Male | 68 (46%) | 39 (60%) | 34 (57%) |
| Female | 79 (54%) | 26 (40%) | 26 (43%) |
| **Infant Ethnicity - Race** |  |  |  |
| Non-White | 63 (43%) | 27 (42%) | 20 (33%) |
| White | 84 (57%) | 38 (58%) | 40 (67%) |
| **Delivery Mode** |  |  |  |
| Vaginal Delivery | 110 (75%) | 48 (74%) | 47 (78%) |
| C-Section | 37 (25%) | 17 (26%) | 13 (22%) |
| **Gestational Age (weeks)** | 39.55 ± 1.17 | 39.71 ± 1.15 | 39.63 ± 1.04 |
| **Birth Weight (Z - score)** | 0.063 ± 0.79 | -0.027 ± 0.86 | -0.0090 ± 0.84 |
| **Infant Feeding Type** |  |  |  |
| Combination |  | 10 (15%) |  |
| Exclusive BF |  | 30 (46%) |  |
| Formula |  | 25 (38%) |  |
| Still BF |  |  | 27 (45%) |
| Stopped BF |  |  | 33 (55%) |
| **Antibiotics (before visit)** |  |  |  |
| FALSE | 134 (100%) | 61 (98%) | 52 (88%) |
| TRUE | 0 (0%) | 1 (1.6%) | 7 (12%) |
| Unknown | 13 | 3 | 1 |
| **Maternal Parity** | 107 (73%) | 44 (68%) | 40 (67%) |
| **Maternal Ethnicity - Race** |  |  |  |
| White | 87 (59%) | 39 (60%) | 40 (67%) |
| Non-White | 60 (41%) | 26 (40%) | 20 (33%) |
| **Maternal Use of Childcare** |  |  |  |
| No |  | 24 (38%) |  |
| Yes |  | 39 (62%) |  |
| **Infant Age at First Dose (weeks)** | 9.80 ± 1.33 | 9.80 ± 1.74 | 9.73 ± 1.51 |
| **Product of First Dose** |  |  |  |
| Rotarix | 0 (0%) | 1 (1.5%) | 1 (1.7%) |
| RotaTeq | 147 (100%) | 64 (98%) | 59 (98%) |
| **Rotavirus Vaccine Product** |  |  |  |
| Combination | 2 (1.4%) | 3 (4.6%) | 1 (1.7%) |
| RotaTeq Vax | 145 (99%) | 62 (95%) | 59 (98%) |
| **Time between Last Dose and Sample Collection (days)** |  | -45.92 ± 35.75 | -201.48 ± 48.08 |
| **Full Vaccination Status** |  |  |  |
| Full Schedule |  | 30 (46%) | 51 (85%) |
| Partial Schedule |  | 35 (54%) | 9 (15%) |

<sup>1</sup>n (%); Mean ± SD
<sup>2</sup> negative value represents days before sample collection

**Figure 1.**
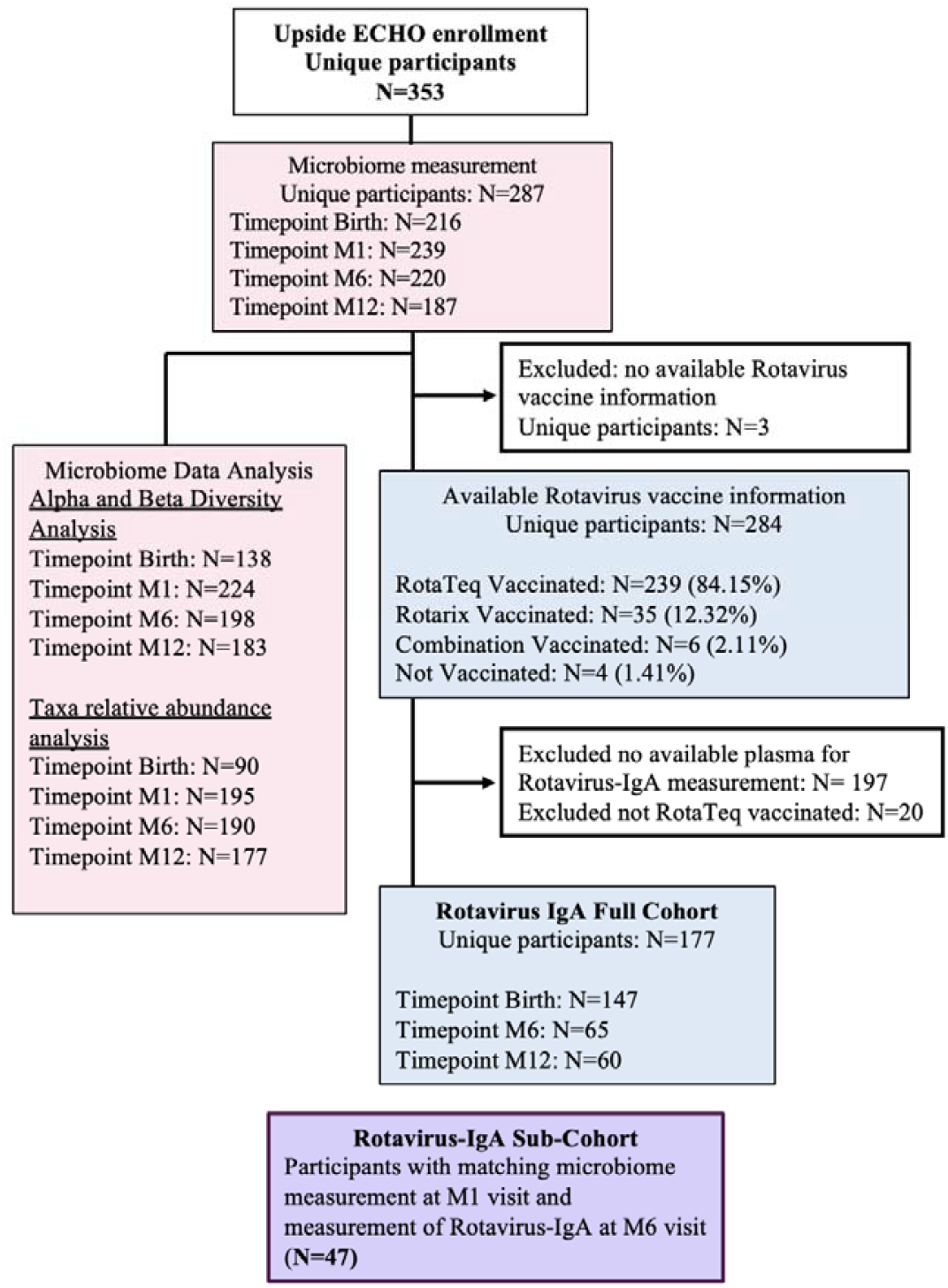
Flow chart of infant participants included in the analyses. Pink boxes indicate the 16S rRNA microbiome arm; blue boxes indicate the rotavirus-IgA arm. Infants with matched microbiome data at month one (M1) and Rotavirus-IgA measurement at month six (M6) are shown in purple.

At the M6 visit (n=65), among infants with available data, 98% had not received antibiotics prior to the M6 visit. By the M6 visit, 46% of participants were fully vaccinated, and 98.5% participants had received RotaTeq for their first dose with a mean age at administration of 9.80 ± 1.74 weeks. Participants received their most recent dose an average of 45.92 ± 35.75 days before the M6 visit (**Table 1**). Prior to the M12 visit (n=60), 88% participants had not received antibiotics. By the M12 visit, 85% of infants were fully vaccinated, with their most recent dose administered a mean of 201.48 ± 48.08 days before the visit, and 98.3% having received the RotaTeq vaccine exclusively (**Table 1**).

### Microbiome 16S rRNA analysis and bacterial taxa comparisons over time

In our population, a significant increase in alpha diversity was observed from birth to the M1 visit (p < 0.0001), from the M1 to the M6 visit (p < 0.0001), and from the M6 to the M12 visit (p < 0.0001) (**Figure 2A**). The changes in beta diversity over time decreased, as shown by the decrease in the distance between the centroids (**Figure 2B**). However, these changes were not statistically significant by PERMANOVA analysis (R^2^ = 0.0047, p = 0.139). We examined the longitudinal changes in mean relative abundance of specific microbial taxa from birth to month twelve (**Figure 2C**). Bacterial genera were grouped by phylum: Actinobacteriota, Bacteroidota, Firmicutes, and Proteobacteria. At birth, Proteobacteria genera, including *Escherichia*-*Shigella*, were the most abundant. By the M1 visit, Proteobacteria decreased in relative abundance over time, and genera from Firmicutes, including *Streptococcus* and *Staphylococcus*, and Actinobacteriota, including *Bifidobacterium*, became more prominent. These patterns remained relatively stable from M6 to M12. Over time, the relative abundance of Bacteroidota genera increased, including *Bacteroides* and *Prevotella*. Between visits M1 and M12, the proportion of genera classified as “Other” also increased within Actinobacteriota and Firmicutes. A comprehensive list of the genera aggregated to the “Other” categories can be found in **Supplemental Table 3** (**appendix p 15-19**).

**Figure 2.**
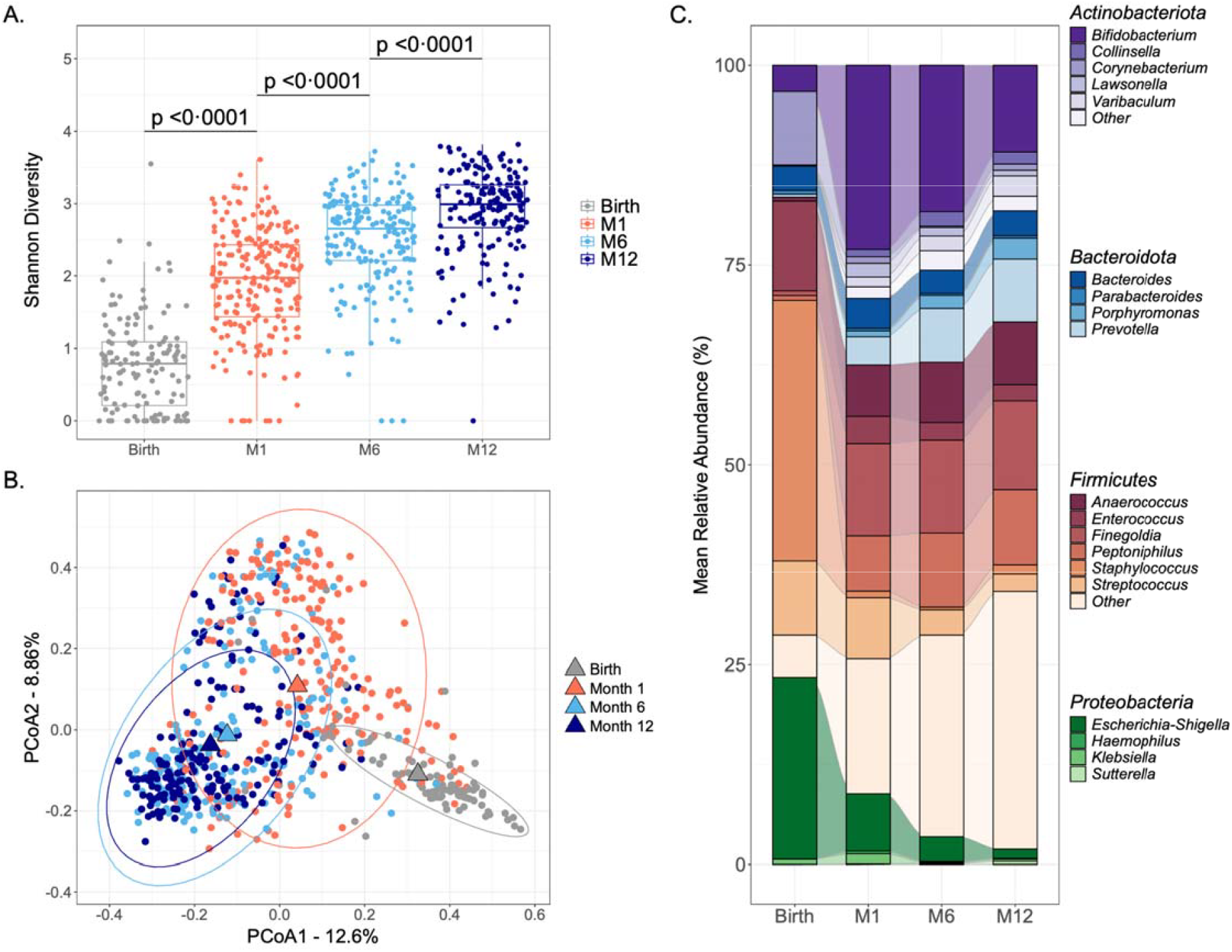
Temporal trends of the infant gut microbiome. Birth (N = 216), Month one (N = 239), Month six (N = 220), and Month 12 (N = 187). (A) Shannon diversity index across visits. P-values from t-tests indicate significant differences between visit groups. (B) Principal Coordinates Analysis (PCoA) of Bray-Curtis dissimilarity by visit. Centroids represent group means, and ellipses show each group’s 95% confidence interval location. (C) Alluvial plot represents bacterial genera mean relative abundance (%) (grouped by Phylum) across visits. Stacked bars represent genus-level composition. Taxa assigned to “Other” categories can be found in **Supplemental Table 3**.

### Sex, ethnicity, breastfeeding, and socioeconomic status influence the infant gut microbiome

Assessment of the relationship between clinical covariates and the developing infant gut microbiome revealed that determinants of the gut microbiome varied across timepoints (**Table 2**). At birth, alpha diversity was positively associated with infant sex (p=0.002) and negatively with mother’s ethnicity-race (p=0.03). At the M1 visit, a positive association with alpha diversity was found with infant sex (p=0.012), enrollment in Medicaid during pregnancy (p=0.016), and mother’s pre-pregnancy BMI (p=0.036). An association between the infant’s alpha diversity and mother’s education, with significant differences observed when the mother had a bachelor’s degree (p=0.016) or some college education (p=0.036). A significant negative association with alpha diversity was found with breastfeeding duration at M1 visit (%) (p<0.0001), a significant positive association with exposure to breastmilk (yes/no) (p<0.0001), as well as marked differences due to feeding type. Compared with exclusively breastfed infants, those who were formula-fed had significantly higher alpha diversity (p < 0.0001), whereas combination-fed infants did not differ from exclusively breastfed infants (p = 0.107).

At the M6 visit, alpha diversity associated positively with Medicaid enrollment during pregnancy (p=0.016) and negatively in mothers with a post-graduate degree (p=0.033). Diet-related variables had the greatest effect on alpha diversity, showing a positive association with breastfeeding exposure (yes/no) and duration at M1 and M6 (%) (p<0.0001), and this effect was sustained at M12 for those who reported continuation of breastfeeding compared to not (p=0.02). Infants who were formula fed (p<0.0001) and combination fed (p=0.007) had significantly higher alpha diversity than exclusively breastfed infants. The M12 visit revealed that females had higher alpha diversity compared to males (p=0.010) (**Table 2**).

**Table 2.** Predictors of the microbiome’s alpha diversity across the first year of life.

| Predictor | Estimate | 95% CI | p-value |
| --- | --- | --- | --- |
| **Birth** |  |  |  |
| **Infant Sex**<br>(Female vs. Male)<sup>1</sup> | 0.33 | [0.12, 0.55] | 0.0025 |
| **Mother's ethnicity-race**<br>(Non-White vs. White)<sup>1</sup> | -0.27 | [-0.52, -0.03] | 0.031 |
| **Month 1** |  |  |  |
| **Infant sex**<br>(Female vs. Male)<sup>1</sup> | 0.24 | [0.05, 0.42] | 0.012 |
| **Breastfeeding duration at M1 (%)** | -0.01 | [-0.01, 0] | <0.0001 |
| **Exposed to BM**<br>(No vs. Yes)<sup>1</sup> | 0.6 | [0.38, 0.83] | <0.0001 |
| **Feeding Type** |  |  |  |
| (Combination vs. Exclusive BF)<sup>1</sup> | 0.2 | [-0.04, 0.45] | 0.11 |
| (Formula vs. Exclusive BF)<sup>1</sup> | 0.65 | [0.42, 0.89] | <0.0001 |
| **Mother's highest education** |  |  |  |
| (<HS or HS vs. Post graduate degree)<sup>1</sup> | 0.24 | [-0.03, 0.53] | 0.087 |
| (Bachelors vs. Post graduate degree)<sup>1</sup> | 0.30 | [0.06, 0.55] | 0.017 |
| (Some college vs. Post graduate degree)<sup>1</sup> | 0.32 | [0.03, 0.62] | 0.034 |
| **Enrolled in Medicaid during Pregnancy**<br>(Yes vs. No)<sup>1</sup> | 0.26 | [0.05, 0.47] | 0.016 |
| **Mother's pre-pregnancy BMI** | 0.01 | [0, 0.03] | 0.036 |
| **Month 6** |  |  |  |
| **Breastfeeding duration at M6 (%)** | -0.005 | [-0.01, 0] | <0.0001 |
| **Exposed to BM**<br>(No vs. Yes)<sup>1</sup> | 0.41 | [0.22, 0.58] | <0.0001 |
| **Feeding Type** |  |  |  |
| (Combination vs. Exclusive BF)<sup>1</sup> | 0.35 | [0.1, 0.58] | 0.0052 |
| (Formula vs. Exclusive BF)<sup>1</sup> | 0.51 | [0.3, 0.69] | <0.0001 |
| **Mother's highest education:** |  |  |  |
| (Bachelors vs. <HS or HS)<sup>1</sup> | -0.22 | [-0.46, 0.03] | 0.089 |
| (Post graduate degree vs. <HS or HS)<sup>1</sup> | -0.28 | [-0.54, -0.02] | 0.033 |
| (Some college vs. <HS or HS)<sup>1</sup> | -0.12 | [-0.4, 0.16] | 0.40 |
| **Enrolled in Medicaid during Pregnancy**<br>(*Yes vs. No*)<sup>1</sup> | 0.31 | [0.12, 0.49] | 0.0010 |
| **Month 12** |  |  |  |
| **Infant Sex**<br>(*Female vs. Male*)<sup>1</sup> | 0.22 | [0.06, 0.39] | 0.010 |
| **Feeding Type**<br>(*Still Breastfeeding vs. Not Breastfeeding*)<sup>1</sup> | -0.21 | [-0.38, -0.03] | 0.020 |

Abbreviations: Breastfeeding (BF); Breastmilk (BM); Body Mass Index (BMI); High school (HS); Less than Highschool (<HS)
Linear regression models were adjusted for covariates, post-conception age, and feeding type.
Only statistically significant variables are shown. Full analysis can be found in **Supplemental Table 2**.
<sup>1</sup>comparison group vs. reference group

### ORV immunogenicity in a healthy US cohort

Of the 65 subjects who received the ORV vaccine, 46% (30/65) received all scheduled doses at the time of sample collection (**Table 1**). Vaccinated infants in our cohort received their first ORV dose by 9.80±1.74 weeks of age. On average, the last dose was received 45.92 ± 35.75 days before Rotavirus-IgA was measured at the M6 visit **(Table 1)**. The proportion of infants who were Rotavirus-IgA positive at birth, indicative of recent (maternal) rotavirus infection, was 3 of 44 (7%) (**Table 3**). A significant rise in antibody from birth to M6 (GMT of 455, 95% CI: [272-761]) was observed (p= 0.045). At the M6 visit, 58 of 65 (89.2%) infants were Rotavirus-IgA seropositive, while 37 of 44 (84.1%) qualified as Rotavirus-IgA seroconverters. Rotavirus-IgA titers decreased at M12 (GMT of 164, 95% CI: [88–305]) compared to the M6 visit (p=0.082) (**Table 3**). At the M6 visit, infants who received the full RotaTeq vaccine schedule (three doses) had higher Rotavirus-IgA titers (GMT = 508, 95% CI: [233–1107]) than those with a partial schedule (one or two doses) (GMT = 414, 95% CI: [202–850]). However, this difference was not statistically significant (p = 0.960) (**Table 4**). The effect of potential correlates of Rotavirus-IgA in our population was tested through linear regression models, shown in **Table 5**. Notably, infants born to Non-White mothers had a 0.346-fold lower Rotavirus-IgA titer compared to those born to White mothers (GMTs: 245, 95% CI: [105-569] vs. 687, 95% CI: [360-1,310], p=0.04).

**Table 3.**
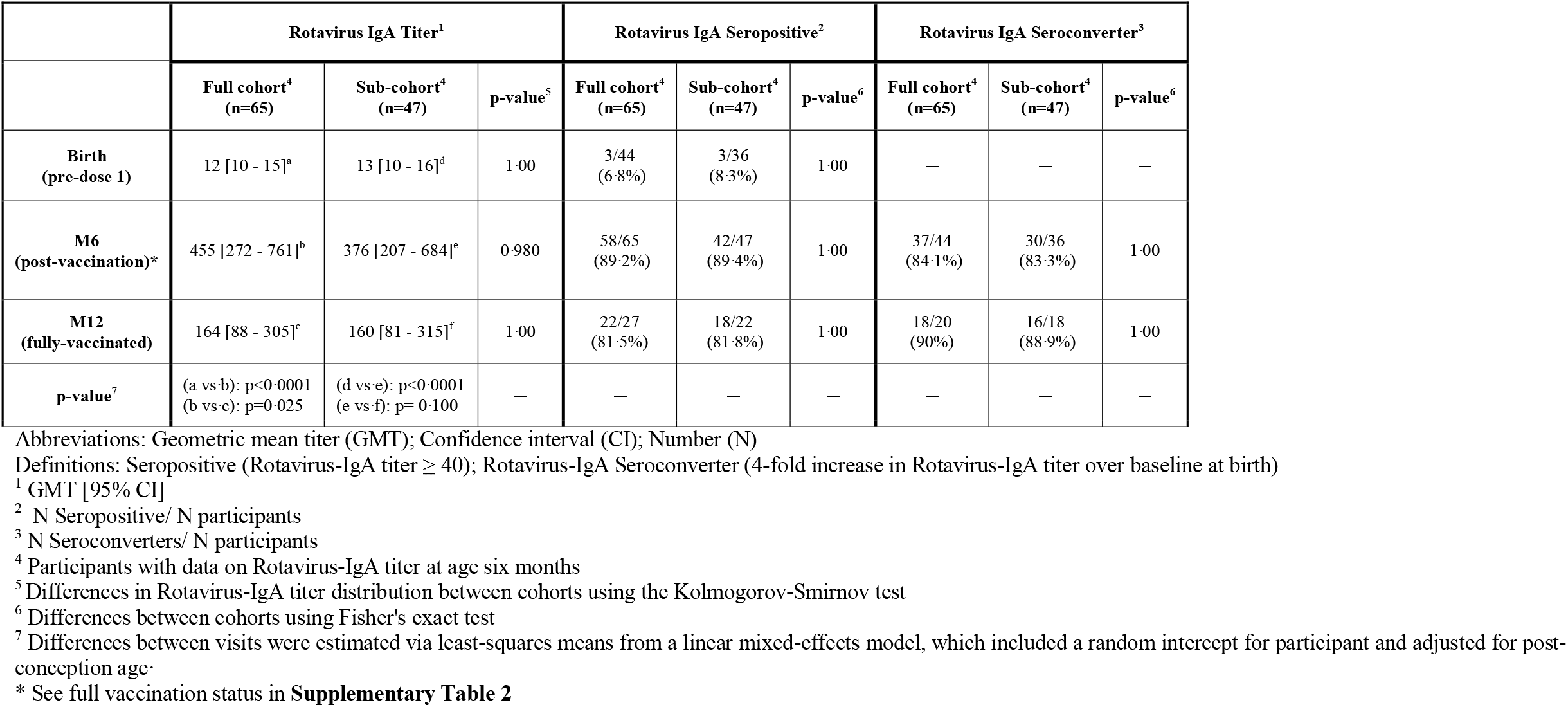
Rotavirus-IgA responses by cohorts.

**Table 4.** Geometric mean titer of vaccination schedule at the M6 visit by cohort.

| **Vaccination schedule status**<br>**at M6 visit** | **Full cohort<sup>1</sup>**<br>**N=65** | **Sub cohort<sup>1</sup>**<br>**N=47** |
| --- | --- | --- |
| Full schedule | 508 [233 - 1107] | 437 [171 - 1117] |
| Partial schedule | 414 [202 - 850] | 337 [147 - 769] |
| p-value<sup>2</sup> | p = 0.960 | p = 0.881 |

Abbreviations: Geometric mean titer (GMT); Confidence interval (CI)
<sup>1</sup> GMT [95% CI]
<sup>2</sup> Linear regression was used to evaluate the association between Rotavirus IgA titer and vaccination schedule status for each cohort.

**Table 5.**
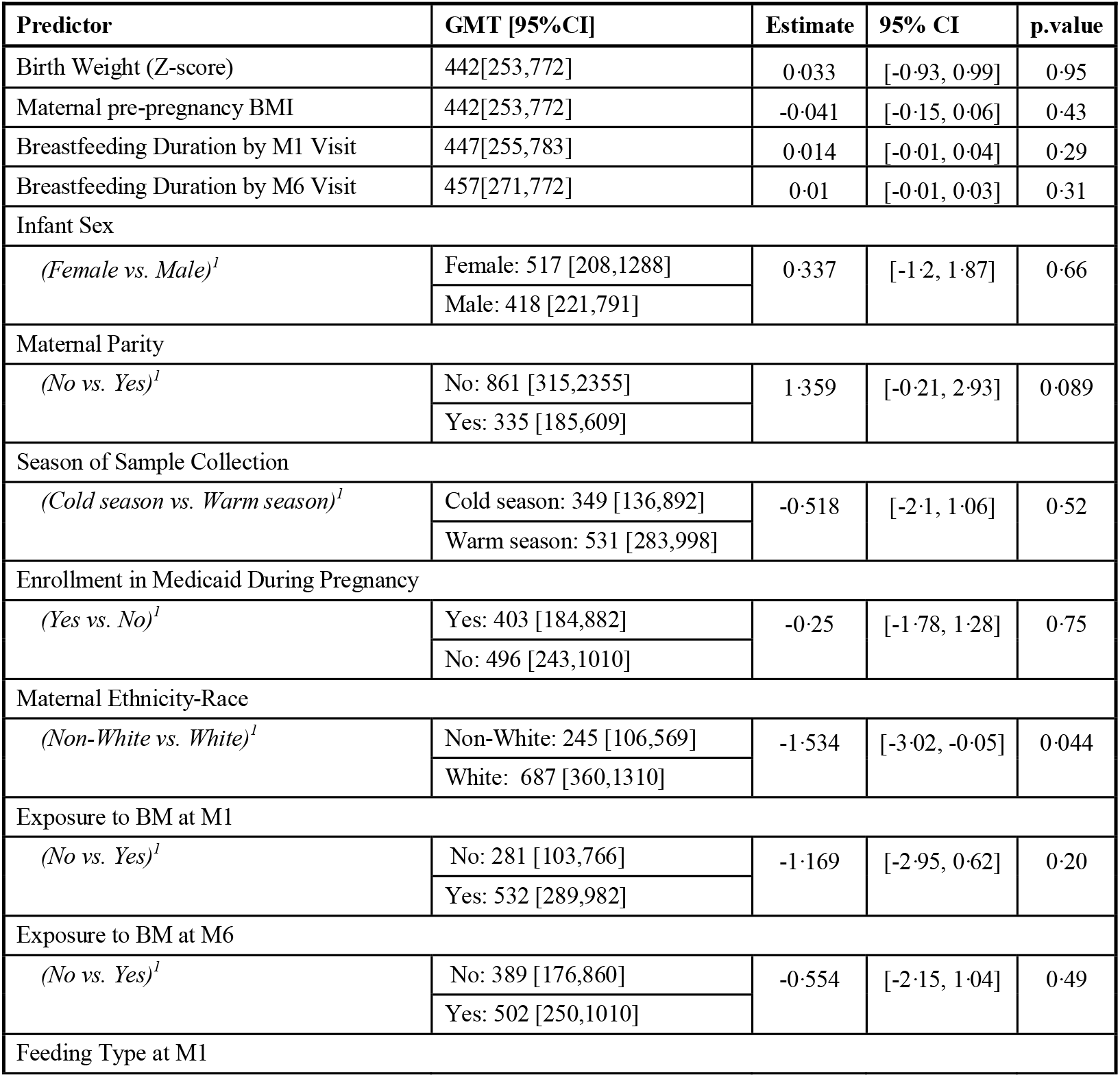
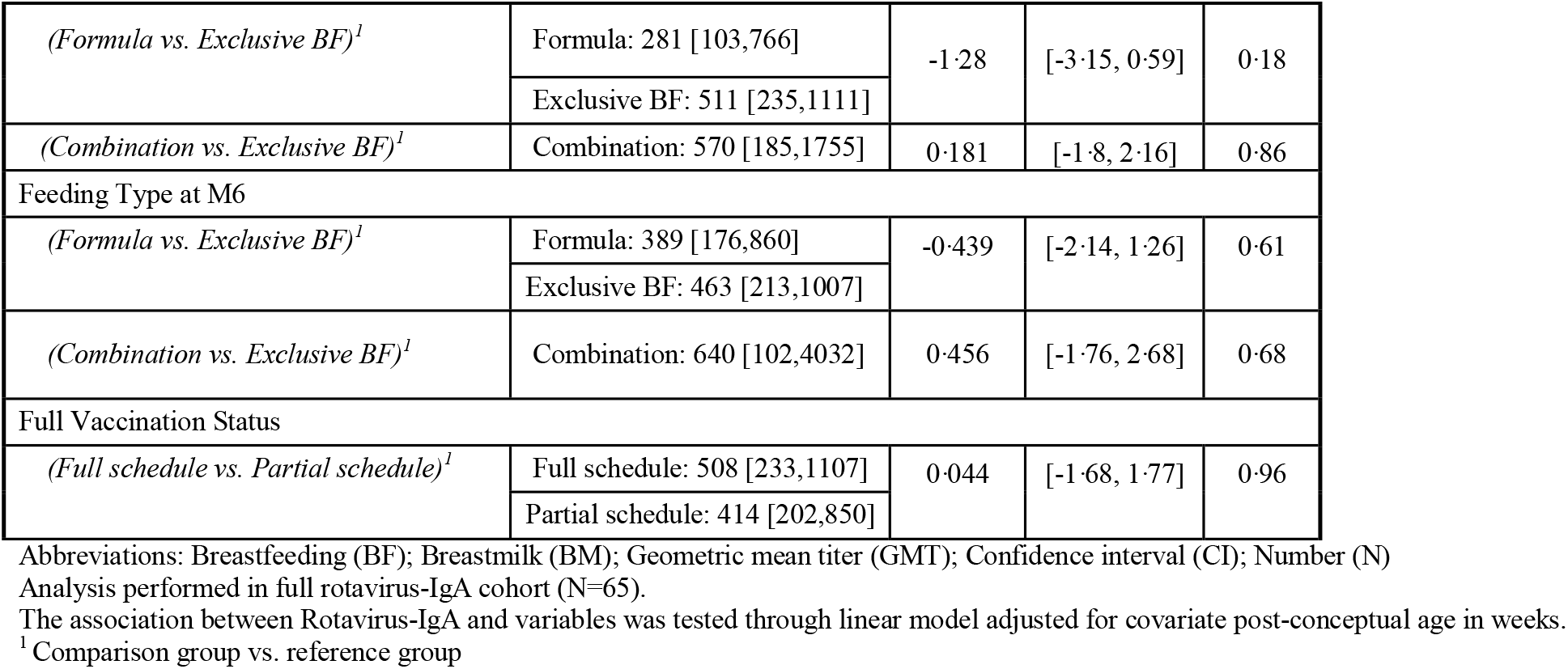
Predictors of Rotavirus-IgA at M6 visit.

| Predictor | GMT [95%CI] | Estimate | 95% CI | p.value |
| --- | --- | --- | --- | --- |
| Birth Weight (Z-score) | 442[253,772] | 0.033 | [-0.93, 0.99] | 0.95 |
| Maternal pre-pregnancy BMI | 442[253,772] | -0.041 | [-0.15, 0.06] | 0.43 |
| Breastfeeding Duration by M1 Visit | 447[255,783] | 0.014 | [-0.01, 0.04] | 0.29 |
| Breastfeeding Duration by M6 Visit | 457[271,772] | 0.01 | [-0.01, 0.03] | 0.31 |
| Infant Sex |  |  |  |  |
| (Female vs. Male)<sup>†</sup> | Female: 517 [208,1288] | 0.337 | [-1.2, 1.87] | 0.66 |
|  | Male: 418 [221,791] |  |  |  |
| Maternal Parity |  |  |  |  |
| (No vs. Yes)<sup>†</sup> | No: 861 [315,2355] | 1.359 | [-0.21, 2.93] | 0.089 |
|  | Yes: 335 [185,609] |  |  |  |
| Season of Sample Collection |  |  |  |  |
| (Cold season vs. Warm season)<sup>†</sup> | Cold season: 349 [136,892] | -0.518 | [-2.1, 1.06] | 0.52 |
|  | Warm season: 531 [283,998] |  |  |  |
| Enrollment in Medicaid During Pregnancy |  |  |  |  |
| (Yes vs. No)<sup>†</sup> | Yes: 403 [184,882] | -0.25 | [-1.78, 1.28] | 0.75 |
|  | No: 496 [243,1010] |  |  |  |
| Maternal Ethnicity-Race |  |  |  |  |
| (Non-White vs. White)<sup>†</sup> | Non-White: 245 [106,569] | -1.534 | [-3.02, -0.05] | 0.044 |
|  | White: 687 [360,1310] |  |  |  |
| Exposure to BM at M1 |  |  |  |  |
| (No vs. Yes)<sup>†</sup> | No: 281 [103,766] | -1.169 | [-2.95, 0.62] | 0.20 |
|  | Yes: 532 [289,982] |  |  |  |
| Exposure to BM at M6 |  |  |  |  |
| (No vs. Yes)<sup>†</sup> | No: 389 [176,860] | -0.554 | [-2.15, 1.04] | 0.49 |
|  | Yes: 502 [250,1010] |  |  |  |
| Feeding Type at M1 |  |  |  |  |
| *(Formula vs. Exclusive BF)<sup>1</sup>* | Formula: 281 [103,766] | -1.28 | [-3.15, 0.59] | 0.18 |
|  | Exclusive BF: 511 [235,1111] |  |  |  |
| *(Combination vs. Exclusive BF)<sup>1</sup>* | Combination: 570 [185,1755] | 0.181 | [-1.8, 2.16] | 0.86 |
| Feeding Type at M6 |  |  |  |  |
| *(Formula vs. Exclusive BF)<sup>1</sup>* | Formula: 389 [176,860] | -0.439 | [-2.14, 1.26] | 0.61 |
|  | Exclusive BF: 463 [213,1007] |  |  |  |
| *(Combination vs. Exclusive BF)<sup>1</sup>* | Combination: 640 [102,4032] | 0.456 | [-1.76, 2.68] | 0.68 |
| Full Vaccination Status |  |  |  |  |
| *(Full schedule vs. Partial schedule)<sup>1</sup>* | Full schedule: 508 [233,1107] | 0.044 | [-1.68, 1.77] | 0.96 |
|  | Partial schedule: 414 [202,850] |  |  |  |

Abbreviations: Breastfeeding (BF); Breastmilk (BM); Geometric mean titer (GMT); Confidence interval (CI); Number (N)
Analysis performed in full rotavirus-IgA cohort (N=65).
The association between Rotavirus-IgA and variables was tested through linear model adjusted for covariate post-conceptual age in weeks.
<sup>1</sup> Comparison group vs. reference group

Additionally, the feeding type and breastmilk duration, whether assessed at the M1 or M6 visit, did not significantly impact Rotavirus-IgA levels in our study population (**Table 5**).

### Rotavirus vaccine response in association with alpha and beta diversity

The M1 microbiome captures the baseline gut ecosystem closest to the time of vaccine initiation, which is important for understanding its foundational influence on immune system priming. Infants who had microbiome data at M1 and Rotavirus-IgA at M6 visit (n=47) were included in this analysis. To ensure generalizability, demographics were compared between the full Rotavirus-IgA cohort and the Sub-cohort (**Supplemental Table 4; appendix p 20**). No differences in either alpha diversity or beta diversity between the full Rotavirus-IgA cohort and the sub-cohort were observed (**Supplemental Figure 1; appendix p 21)**. On average, participants received their first ORV dose 27.41± 13.18 days after collecting the M1 visit rectal swab sample used for microbiome sequencing (**Supplemental Table 4; appendix p 20**). We found each one-unit increase in alpha diversity at M1 visit was linked to an estimated 4.45-fold increase in Rotavirus-IgA titers, accounting for approximately 10.8% of the variance observed (ß= 2.151, 95% CI:[0.31-3.99], p=0.023) (**Figure 3A**). In contrast, no evidence of an association between the overall microbial composition at M1 and Rotavirus-IgA responses was found (p = 0.937) (**Figure 3B**).

**Figure 3.**
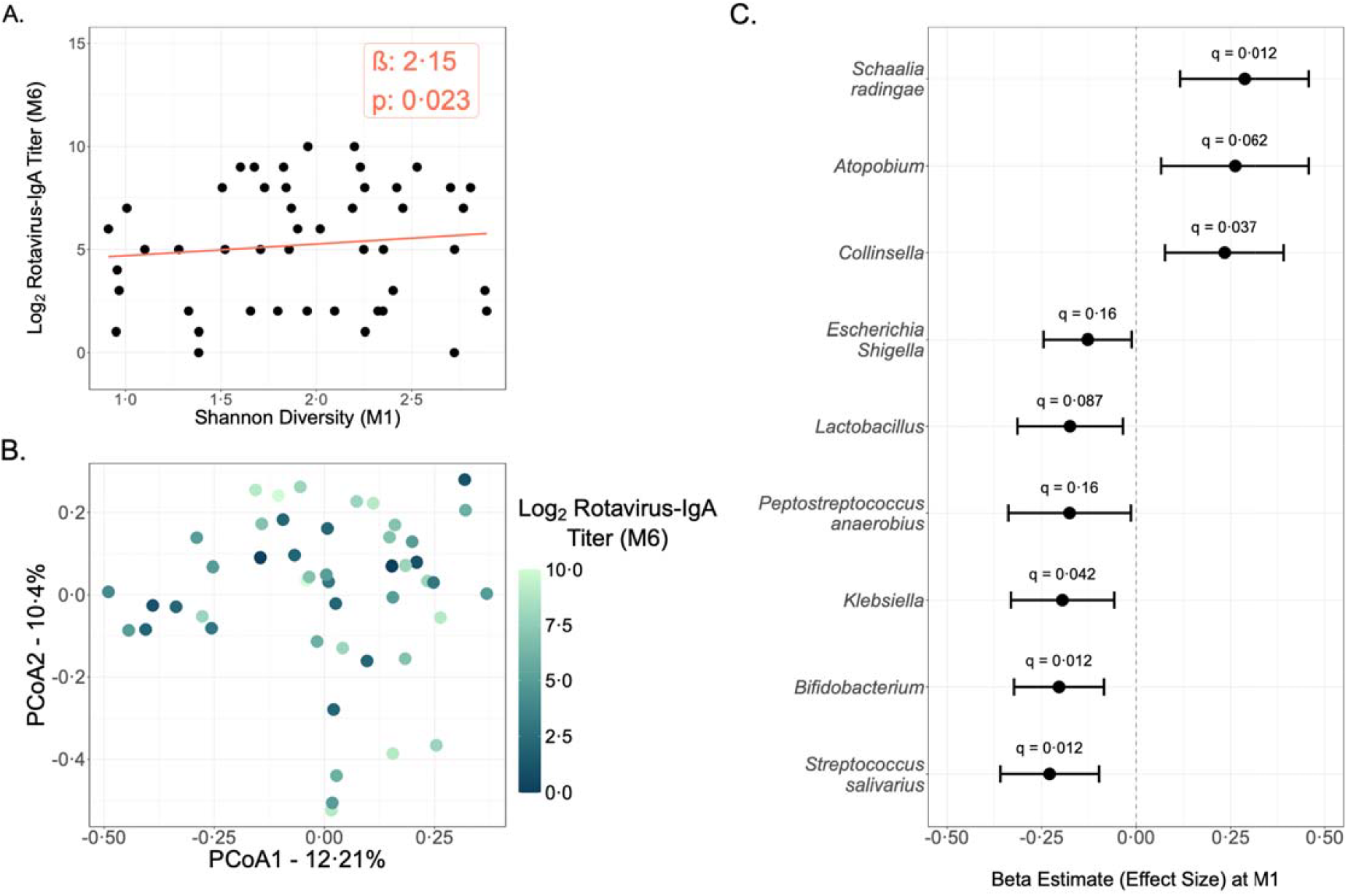
Association between the infant gut microbiome at month one and Rotavirus-IgA at M6 visit. (A) Shannon diversity at M1 visit (N = 47) by Rotavirus-IgA titers. The box represents linear regression results. Shannon’s diversity association was tested through a linear regression model accounting for covariates. (B) Beta diversity at month one. Overlay color represents the participant’s corresponding Rotavirus-IgA titer at month six. Beta diversity variance explained by Rotavirus-IgA titers was tested through a microbiome regression-based kernel association test. (C) Forest plot illustrating the association between taxa at month one and Rotavirus-IgA. Points indicate the Beta Estimate for each association, with bars representing the corresponding 95% Confidence Interval (CI). A lasso-penalized log contrast model was used to assess the association between microbial taxa and RV-IgA titer, adjusting for the post-conception age of both microbiome and plasma samples. P-values were corrected for multiple comparisons using the Benjamini-Hochberg FDR method.

### Microbial taxa associated with vaccine response

We performed an L1 penalized log-contrast regression to assess the associations between individual taxa at M1 and Rotavirus-IgA at M6, while adjusting for the post-conception ages of the microbiome and plasma at collection (**Figure 3C**). Our analysis revealed Rotavirus-IgA titers significantly associated with multiple taxa: positively with *Collinsella* (ß: 0.243, 95% CI:[0.076, 0.392], q= 0.037), *Atopobium* (ß: 0.262, 95% CI:[0.066, 0.458], q= 0.062), and *Schaalia radingae* (ß: 0.28, 95% CI:[0.116, 0.458], q=0.018); and negatively with *Bifidobacteria* (ß: −0.204, 95% CI:[−0.323, −0.085], q= 0.012), *Klebsiella* (ß: −0.195, 95% CI:[−0.331, −0.058], q= 0.042), *Lactobacillus* (ß: −0.17, 95% CI:[−0.314, −0.035], q= 0.087), *Escherichia-Shigella* (ß: −0.128, 95% CI:[−0.245, −0.012], q= 0.162), *Streptococcus salivarus* (ß: −0.229, 95% CI:[−0.359, −0.098], q= 0.012), and *Peptostreptococcus anaerobius* (ß: −0.176, 95% CI:[−0.338, −0.014], q= 0.162).

## Discussion

Vaccines, including ORVs, elicit variable immune responses across individuals, with particularly pronounced differences among children from LMIC and HIC.^3^ The gut microbiome has emerged as a potential contributor to this variability, especially in early life when microbial colonization and immune system development are tightly intertwined.^23^ Several studies have investigated the microbiome-ORV relationship, but findings have been inconsistent, likely reflecting differences in study design, geography, and host context.^13–20^ To date, all eight published studies have focused on LMICs, where perinatal factors, environmental exposures, nutritional status, and baseline microbial profiles strongly influence both ORV immunogenicity and gut ecology. Yet, understudied HIC cohorts such as in the U.S. with low viral prevalence and high vaccine uptake may offer greater precision when examining microbiome-ORV relationships.

Here, we present the first U.S.-based study to report a significant association between the infant gut microbiome and ORV immune responses. Among infants with high vaccine uptake, Rotavirus-IgA titers varied considerably despite the expected high seroconversion. These observations underscored the value of antibody titers, rather than seroconversion alone, for understanding immune heterogeneity in HIC populations. In our study, the infant gut microbiome was characterized at the M1 visit, a time point preceding the Rotavirus-IgA measurement and close to the time when infants receive their first dose of the ORV. We considered the microbiome structure prior to Rotavirus response measurement as a key factor influencing the generation of a robust vaccine-induced response. We found that higher gut microbial diversity at the M1 was positively associated with ORV-induced immune responses, results which align with a recent multicountry LMIC study that similarly reported positive associations between microbial diversity, IgA seroconversion, and vaccine shedding.^18^ Together, these data strengthen the hypothesis that early-life microbial diversity is a reproducible marker, and potentially a modifiable determinant of ORV immunogenicity.

Specific microbial taxa were also significantly associated with downstream Rotavirus-IgA titers. *Collinsella, Atopobium*, and *Schaalia radingae* were positively associated with Rotavirus-IgA titers. *Collinsella* has been linked to IFN-γ□ CD8□ T cell responses in a Rotavirus infection model^24^ and to increased intestinal permeability and Il-17A induction.^25^ These features of mucosal immune activation, particularly Th17 responses, are known to support differentiation of IgA-secreting plasma cells^26^, providing a plausible mechanistic link between the microbe and enhanced Rotavirus-IgA production. More broadly, bacterial taxa may influence ORV immunogenicity via the secretion of bacteria-derived adjuvants such as flagellin or lipopolysaccharide, or via microbial metabolic products like short-chain fatty acids, which have been shown to enhance IgA class switching.^27^ Granular metagenomics and metabolomics assessment could further inform mechanisms by which these taxa support ORV immunogenicity and IgA production.

Conversely, *Bifidobacterium, Lactobacillus, Klebsiella, Escherichia-Shigella, Streptococcus salivarius*, and *Peptostreptococcus anaerobiu*s were negatively associated with Rotavirus-IgA titers. While *Lactobacillus* and *Escherichia* have been linked to protection against rotavirus disease^28^, they may undermine vaccine-induced immunity by fostering conditions that limit viral replication and dampen pro-inflammatory signaling. Our findings are consistent with those reported by Wagner and colleagues, who observed increased abundance of *Streptococcus, Klebsiella*, and Enterobacteriaceae, impaired IgA seroconversion, and reduced vaccine shedding in infants from Malawi and Indonesia.^18^ However, they differ from Harris and colleagues, who reported positive correlations between *Escherichia* and Rotavirus-IgA titers in Ghana.^17^ Such discrepancies may reflect geographic and immunological context-specific effects, highlighting the complexity of host-microbiome-vaccine interactions and the importance of studying population-specific effects.

Breastfeeding practices shape the infant gut microbiome and represent a potential confounder in analyses of microbiome-immune interactions. Yet they are often overlooked in studies, including those investigating ORV immunogenicity. Evidence supporting a direct relationship between breastfeeding and ORV immunogenicity is inconsistent across populations.^14,29^ Consistent with other HIC-based studies, our analysis revealed no relationship between breastfeeding variables and Rotavirus-IgA titers. ^29^ Our results do suggest that while breastfeeding is a crucial factor in early microbiome development, its impact on ORV immunogenicity in HIC populations is likely minimal. We also observed lower Rotavirus-IgA responses in infants born to non-white mothers compared to those from white mothers. One potential explanation is maternal FUT2 secretor status, known to influence both rotavirus susceptibility and ORV immunogenicity.^29^ However, FUT2 non-secretor prevalence is relatively consistent across racial and ethnic groups, affecting approximately 20% of the population.^30^ Thus, the observed disparity in differences in Rotavirus-IgA titers is unlikely to be explained by secretor status alone. Rather, these findings are likely to reflect a more intricate interplay of genetic, immunological, and environmental factors.

A key strength of our study lies in its controlled design and relatively homogeneous cohort, allowing us to minimize biological and environmental confounders that can bias microbiome-immune associations. By focusing on healthy infants of similar demographic backgrounds who uniformly received the RotaTeq vaccine, we improved internal validity despite the modest sample size. Nevertheless, we were unable to account for maternal IgG and FUT2 secretor status, known to influence susceptibility to rotavirus infection and vaccine response in populations similar to ours.^29^ Additionally, our analysis was restricted to bacterial composition and a single immune endpoint. Expanding to include other ORV immune outcomes and applying metagenomic and metabolomic approaches will be essential to define mechanisms.

In conclusion, this study highlights the critical window of early-life microbial exposure priming mucosal immunity. We show a strong association between the gut microbiome pre-vaccination and ORV Rotavirus-IgA titers post-vaccination. By linking early-life microbiome diversity and composition to ORV antibody responses in U.S. infants, this study establishes the microbiome as a critical determinant of ORV immunogenicity in HICs. Continued investigation through large, longitudinal, and multi-omic studies will be key to unlocking actionable insights for optimizing ORV immunogenicity and efficacy across populations.

## Data Availability

All data produced in the present study are available upon reasonable request to the authors.

## Data Sharing

Illumina 16S rRNA V3V4 amplicon sequences will be deposited and available in the Sequence Read Archive upon publication.

## Acknowledgements

The authors of this study would like to acknowledge the support of the mothers and infants participating in UPSIDE-ECHO study, UPSIDE-ECHO staff, the Babies, Bugs, and Brains team, Scheible lab members, Dr. Darline Castro-Melendez, Dr. Adam Geber, and Dr. Robert López-Astacio. We would like to acknowledge the funding sources of this study, NIH (UPSIDE-ECHO) 5UH3OD023349, NIH (Babies, Bugs and Brains) R01-MH125103, and the NIH (U of R CTSA) TL1 TR002000.

