## Supplemental Materials for "Early-life gut microbiome is associated with immune response to the oral rotavirus vaccine in healthy infants in the US"

### Supplemental Methods

#### DNA Extraction and 16S rRNA amplicon sequencing

Total genomic DNA was extracted from rectal samples by modifying the Quick-DNA™ Fecal/Soil Microbe Miniprep Kit (Zymo Research; Irvine, CA). Samples were then homogenized with bashing beads to break open the cells using FastPrep mechanical lysis (MPBio, Solon, OH). The V3-V4 hypervariable regions of 16S rRNA were amplified with Phusion High-Fidelity polymerase (New England Biolabs, Ipswich, MA) using dual-indexed primers specific to the V3-V4 regions (319F: 5′ ACTCCTACGGGAGGCAGCAG 3′; 806R: 3′ ACTCCTACGGGAGGCAGCAG 5′). (Holm, JB. 2019) Amplicons were then pooled using SequalPrep™ normalization plates (Thermo Fisher Scientific; Waltham, MA). Pooled libraries were then sequenced on an Illumina MiSeq (Illumina; San Diego, CA) at the University of Rochester Genomics Research Center. Each sequencing run included the following controls: 1) positive control containing a 1:5 mixture of *Staphylococcus aureus*, *Lactococcus lactis*, *Porphyromonas gingivalis*, *Streptococcus mutans*, and *Escherichia coli*; 2) negative control consisting of sterile saline; and 3) extraction controls of sterile saline.

#### Taxonomical Classification

Raw read data from the Illumina MiSeq was first converted into a FASTQ format 2x312 paired-end sequenced file using bcl2fastq version 1.8.4, provided by Illumina. Reads were multiplexed using a configuration described previously by Holm JB.[1] The first 24-29 bases in each paired read represented an adapter, followed by an 8-base barcode, an overlapping tag sequence, and a heterogeneity spacer. After that, a gene-specific primer was present, leading to the target 16S rRNA sequence.

Demultiplexed reads were then imported into QIIME2 (version 2022.2) for subsequent analysis.[2] Reads were demultiplexed, requiring exact barcode matches, and 16S primer sequences were removed, allowing 20% mismatched and requiring at least 18 bases. Next, the data were cleaned, joined, and denoised using DADA2. The totality of the infant samples spanned five sequencing runs. For all sequencing runs, forward reads were truncated to 275 bps and reverse reads to 260 bps, error profiles were learned with a sample of one million reads, and a maximum expected error of two was allowed. All batches were then merged before taxonomical classification. The taxonomical classification was performed using a custom Bayesian classifier trained with full-length 16S sequences from the SILVA classifiers. [3,4]

**16S rRNA sequencing data post-processing**

To reduce noise and improve the reliability of the analysis, we implemented a series of cutoffs for both samples and amplicon sequence variants (ASVs)[5]. For diversity analyses, ASVs present in ≤ 2 samples were removed. ASVs at the species level were serially aggregated up to the phylum level, utilizing different cutoffs depending on the analysis to reduce false positives due to excess zeros. For instance, 20% was used as a cutoff for association analyses, e.g., if a taxon at the species level was present in fewer than 20% of samples, it was aggregated into its genus. Taxa not meeting these respective presence thresholds were removed before further analysis. We also removed samples that had less than 100 reads in total. A detailed breakdown of the post-processing can be found in **Supplemental Table 1**.

**Supplemental Table 1 Cutoff used for 16S data analyses.**

| *Analysis* | *Sample read cutoff (reads)* | *Taxonomical aggregation level* | *Taxonomic aggregation percent* | *Taxa removal cutoff* |
|---|---|---|---|---|
| *Alpha and beta diversities* | *5* | — | — | *2 samples* |
| *Sub-composition of all timepoints* | *115* | *Up to Genus* | — | *10% samples* |
| *Log contrast model* | *100* | *Up to Phylum* | *20%* | *20% samples* |

**Rotavirus-IgA ELISA**

Rotavirus-IgA in plasma was measured utilizing an Enzyme-Linked Immunoassay (EIA)[6]. In brief, 96-well microplates were coated with 100 µl/well of rabbit hyperimmune serum to the rhesus rotavirus strain (RRV) (Source: CDC) at a 1:10,000 dilution and then incubated overnight. The microplates were incubated with 200µL Blocking Buffer (5% wt/vol BD Difco Skim milk in PBS) at 37ºC for 1 hour. Microplates were then incubated for 1 hour in duplicate with 100 µl/well of clarified lysates of RotaTeq G1 strain-infected MA104 cell cultures in working dilutions of 7.2 × 10^5 FFU/mL for RotaTeq antigen. A 100 µl/well of serially diluted test plasma (1:20–2:10,240) was added and incubated for 2 hours. Followed by adding 100 µl/well of biotin-conjugated goat anti-human IgA (KPL, USA) diluted 1:2,000 and incubated for 1 hour. Extravidin (100 µl/well) (Sigma, USA) diluted 1:3,000 was added to the wells and incubated for 1 hour, and then the reactions were developed with 100 µl/well 3,30,5,50-tetramethylbenzidine (Sigma, USA) and stopped with 100 µl/well 1 N HCl. Plate optical density was measured using a BioTek 800TS Plate reader set to 450 nm reading reference wavelength. Rotavirus-IgA titers in plasma were calculated as the reciprocal of the highest dilution that gave a mean OD greater than the cutoff value (3 standard deviations above the mean OD of the negative control serum wells).

**Rotavirus-IgA titer transformation and GMT calculation**

The primary immunological outcome of this study was the measurement of Rotavirus-IgA titers. The Rotavirus-IgA titers were recorded as a $y = \log_2(\text{dilution}/10)$. A one-unit increase corresponds to a two-fold change in the raw titer. This approach facilitated a more biologically interpretable understanding of variations in antibody concentration. The geometric mean titers (GMT) were defined as $y = 10 * 2^\wedge(mean\ \log_2 titer)$.

**Descriptive Statistics and Univariate Analyses**

Descriptive statistics were used to summarize the characteristics of the study population, providing a comprehensive overview of key variables and patterns. Means and standard deviations were reported for continuous variables, while frequencies and percentages relative to the total number of observations were reported for each level of categorical variables. For between-group comparisons, the Wilcoxon-rank-sum test was performed for continuous variables, and Pearson's Chi-squared test was performed for categorical variables. A p-value less than 0.05 was considered statistically significant. For analyses involving multiple hypotheses, the Benjamini–Hochberg procedure was applied to control the false discovery rate at 0.05 level. All statistical analyses were conducted in R version 4.5.0 (R Foundation for Statistical Computing, Vienna, Austria).

**Infant Gut Microbiome Statistical Analyses**

To determine which variables influenced the developing gut microbiome in our cohort, we took a cross-sectional approach. We first assessed between-group differences in alpha diversity for the variable of interest using the Wilcoxon rank-sum or the Kruskal-Wallis test for nominal variables and Spearman's rank correlation coefficient for continuous values (**Supplemental Table 2**). As the effect of the variables with significant differences in their Shannon diversity might be confounded by age and/or feeding type, we performed a linear regression analysis with and without age and feeding type.

**Rotavirus Vaccine Immune Outcomes Statistical Analyses**

We modeled longitudinal Rotavirus-IgA responses using an LMER model with month visit as the predictor, gestational age as a covariate, and a random intercept for each participant to account for serial correlation in repeated measures in Rotavirus-IgA responses. The lsmeans were computed for pairwise comparisons of visits. Differences in Rotavirus-IgA titer distribution at birth, M6, and M12 between the full cohort of infants with Rotavirus-IgA measurements (n = 65) and the sub-cohort with both month one microbiome and month six Rotavirus-IgA data (n = 47) were tested using the Kolmogorov-Smirnov test. Differences in seropositivity and seroconversion rates between cohorts at each visit were assessed using Fisher's exact test. A linear regression model was utilized to test the association between vaccination status at the M6 visit and Rotavirus-IgA titers.

To further explore associations between Rotavirus-IgA titers at the M6 visit and individual covariates, we applied univariate analyses using Wilcoxon rank-sum or Kruskal–Wallis tests (for nominal variables such as demographics or feeding practices) and Spearman's rank correlation test (for continuous variables such as breastfeeding duration). We further fitted simple linear regression models for each covariate of interest, adjusting for post-conception age, to obtain the effect estimates and 98% confidence intervals (95%CI).

**Gut Microbiome–Rotavirus-IgA Associations**

The relationship between microbial alpha diversity at the M1 visit and Rotavirus-IgA titers at the M6 visit was assessed using a linear regression model. This model adjusted for relevant covariates, including post-conception age (defined as gestational age plus age at visit) at both the M1 microbiome sampling and the M6 plasma collection, as well as feeding type at the M1 visit. To evaluate the overall association between M1 microbiome composition (beta diversity) and Rotavirus-IgA titers at M6, we applied Microbiome Regression-Based Kernel Association Tests (MiRKAT[7]).

To identify microbial taxa at the M1 visit associated with Rotavirus-IgA at the M6 visit, we employed an $L_1$-penalized log-contrast model followed by a debiasing procedure specifically developed for compositional covariates.[8] Post-conception age was included as a covariate in these models to control for its confounding effects. Due to the multiple testing nature of these analyses, the Benjamini–Hochberg procedure was applied to control the false discovery rate at the 0.05 level. All analyses were conducted in RStudio (version 4.5.0) with the following R packages: *lme4*[9], *emmeans*[10], *rstatix*[11], *vegan*[12] and *MiRKAT*[7] .

**Supplemental Results**

Supplemental Table 2 Univariate analysis of covariates of the infant gut microbiome.

| Timepoint | Variable | Group | mean±sd | group1 | group2 | n1 | n2 | statistic | estimate | 95%CI | p | p·adj[1] |
|---|---|---|---|---|---|---|---|---|---|---|---|---|
| Birth | | | | | | | | | | | | |
| | Age (weeks) | | 0·17 ± 0·10 | | | | | 394556·1775 | 0·079 | | 0·36 | |
| | Post-conception age (weeks) | | 39·96 ± 1·22 | | | | | 401701·8169 | 0·063 | | 0·47 | |
| | Gestational Age | | 39·79 ± 1·22 | | | | | 411087·344 | 0·061 | | 0·47 | |
| | Maternal Pre-Pregnancy BMI | | 27·77 ± 6·90 | | | | | 325705·7159 | -0·001 | | 0·99 | |
| | Weight at Birth (Z-score) | | -0·15 ± 0·86 | | | | | 285696·4015 | 0·122 | | 0·17 | |
| | Infant Sex | Male | 0·62 ± 0·56 | Male | Female | 73 | 65 | 1642·5 | -0·310 | (-0·553,-0·09) | 0·002 | |
| | | Female | 0·96 ± 0·71 | | | | | | | | | |
| | Infant Ethnicity | Non-Hispanic or Non-Latino | 0·82 ± 0·66 | Non-Hispanic or Non-Latino | Hispanic or Latino | 124 | 14 | 1154 | 0·355 | (0, 0·691) | 0·044 | |
| | | Hispanic or Latino | 0·46 ± 0·50 | | | | | | | | | |
| | Infant Ethnicity-Race | White | 0·84 ± 0·64 | Non-White | White | 43 | 95 | 1643·5 | -0·179 | (-0·454, 0) | 0·067 | |
| | | Non-White | 0·65 ± 0·66 | | | | | | | | | |
| | Season of birth | Spring | 0·62 ± 0·45 | Fall | Spring | 44 | 31 | 784 | 0·154 | (-0·088, 0·512) | 0·27 | 0·45 |
| | | Summer | 1·00 ± 0·81 | Fall | Summer | 44 | 32 | 646 | -0·080 | (-0·432, 0·223) | 0·55 | 0·65 |
| | | Fall | 0·84 ± 0·68 | Fall | Winter | 44 | 31 | 778 | 0·102 | (-0·101, 0·504) | 0·30 | 0·45 |
| | | Winter | 0·65 ± 0·54 | Spring | Summer | 31 | 32 | 371·5 | -0·231 | (-0·603, 0·028) | 0·09 | 0·34 |
| | | | | Spring | Winter | 31 | 31 | 467 | 0·000 | (-0·257, 0·226) | 0·85 | 0·85 |

| Timepoint | Variable | Group | mean±sd | group1 | group2 | n1 | n2 | statistic | estimate | 95%CI | p | p·adj[1] |
|---|---|---|---|---|---|---|---|---|---|---|---|---|
| | | | | Summer | Winter | 32 | 31 | 612 | 0·239 | (-0·048, 0·64) | 0·11 | 0·34 |
| | Mode of Delivery | Vaginal Delivery | 0·79 ± 0·61 | Vaginal Delivery | C-Section | 117 | 21 | 1338·5 | 0·069 | (-0·163, 0·447) | 0·52 | |
| | | C-Section | 0·76 ± 0·86 | | | | | | | | | |
| | Enrolled in Medicaid during Pregnancy | Yes | 0·67 ± 0·67 | No | Yes | 87 | 51 | 2642 | 0·161 | (0, 0·416) | 0·062 | |
| | | No | 0·85 ± 0·64 | | | | | | | | | |
| | Maternal Ethnicity-Race | White | 0·87 ± 0·63 | Non-White | White | 42 | 96 | 1415 | -0·293 | (-0·565, -0·058) | 0·005 | |
| | | Non-White | 0·59 ± 0·68 | | | | | | | | | |
| | Maternal Education | Post graduate degree | 0·82 ± 0·66 | Post graduate degree | <HS or HS | 41 | 31 | 645 | 0·000 | (-0·291, 0·337) | 0·92 | 0·99 |
| | | <HS or HS | 0·83 ± 0·76 | Post graduate degree | Some college | 41 | 24 | 568 | 0·111 | (-0·122, 0·457) | 0·30 | 0·71 |
| | | Bachelors | 0·80 ± 0·63 | Post graduate degree | Bachelors | 41 | 42 | 871·5 | 0·000 | (-0·237, 0·278) | 0·93 | 0·99 |
| | | Some college | 0·64 ± 0·54 | <HS or HS | Some college | 31 | 24 | 427 | 0·088 | (-0·174, 0·452) | 0·35 | 0·71 |
| | | | | <HS or HS | Bachelors | 31 | 42 | 649 | 0·000 | (-0·294, 0·29) | 0·99 | 0·99 |
| | | | | Some college | Bachelors | 24 | 42 | 432 | -0·091 | (-0·457, 0·112) | 0·34 | 0·71 |
| | Maternal Parity | Yes | 0·78 ± 0·65 | Yes | No | 94 | 44 | 1989·5 | -0·011 | (-0·209, 0·191) | 0·72 | |
| | | No | 0·79 ± 0·66 | | | | | | | | | |
| M1 | | | | | | | | | | | | |
| | Gestational Age | | 39·61 ± 1·19 | | | | | 2073957·217 | -0·107 | | 0·11 | |
| | Maternal Pre-Pregnancy BMI | | 28·10 ± 7·23 | | | | | 1081472·252 | 0·224 | | 0·0013 | |

| Timepoint | Variable | Group | mean±sd | group1 | group2 | n1 | n2 | statistic | estimate | 95%CI | p | p·adj[1] |
|---|---|---|---|---|---|---|---|---|---|---|---|---|
| | Weight at Birth (Z-score) | | -0·06 ± 0·84 | | | | | 1450211·125 | -0·040 | | 0·57 | |
| | Breastfeeding Duration at M1 (%) | | 86·18 ± 30·26 | | | | | 2485424·129 | -0·327 | | <0·0001 | |
| | Infant Sex | Female | 2·05 ± 0·70 | Male | Female | 116 | 108 | 5197 | -0·218 | (-0·411, -0·019) | 0·028 | |
| | | Male | 1·79 ± 0·76 | | | | | | | | | |
| | Infant Ethnicity | Non-Hispanic or Non-Latino | 1·93 ± 0·71 | Non-Hispanic or Non-Latino | Hispanic or Latino | 200 | 24 | 2680 | 0·190 | (-0·214, 0·58) | 0·35 | |
| | | Hispanic or Latino | 1·76 ± 0·97 | | | | | | | | | |
| | Infant Ethnicity-Race | White | 1·75 ± 0·68 | Non-White | White | 77 | 147 | 7922 | 0·514 | (0·325, 0·707) | <0·0001 | |
| | | Non-White | 2·23 ± 0·75 | | | | | | | | | |
| | Antibiotics | TRUE | 1·75 ± 0·70 | FALSE | TRUE | 217 | 7 | 865 | 0·177 | (-0·39, 0·752) | 0·534 | |
| | | FALSE | 1·92 ± 0·75 | | | | | | | | | |
| | Mode of Delivery | Vaginal Delivery | 1·90 ± 0·76 | Vaginal Delivery | C-Section | 175 | 49 | 4040·5 | -0·075 | (-0·31, 0·162) | 0·539 | |
| | | C-Section | 1·98 ± 0·68 | | | | | | | | | |
| | Enrolled in Medicaid during Pregnancy | No | 1·74 ± 0·69 | No | Yes | 138 | 86 | 3705·5 | -0·469 | (-0·655, -0·29) | <0·0001 | |
| | | Yes | 2·19 ± 0·74 | | | | | | | | | |
| | Maternal Ethnicity-Race | White | 1·82 ± 0·68 | Non-White | White | 77 | 147 | 7145·5 | 0·350 | (0·146, 0·555) | 0·0013 | |
| | | Non-White | 2·10 ± 0·82 | | | | | | | | | |
| | Maternal Education | Post graduate degree | 1·61 ± 0·67 | Post graduate degree | <HS or HS | 72 | 59 | 1232 | -0·522 | (-0·786, -0·297) | <0·0001 | <0·0001 |
| | | <HS or HS | 2·12 ± 0·79 | Post graduate degree | Some college | 72 | 34 | 768·5 | -0·522 | (-0·787, -0·224) | 0·002 | 0·006 |

| Timepoint | Variable | Group | mean±sd | group1 | group2 | n1 | n2 | statistic | estimate | 95%CI | p | p·adj[1] |
|---|---|---|---|---|---|---|---|---|---|---|---|---|
| | | Bachelors | 1·96 ± 0·68 | Post graduate degree | Bachelors | 72 | 59 | 1498 | -0·340 | (-0·577, -0·111) | 0·004 | 0·008 |
| | | Some college | 2·12 ± 0·74 | <HS or HS | Some college | 59 | 34 | 1010 | 0·013 | (-0·299, 0·385) | 0·96 | 0·96 |
| | | | | <HS or HS | Bachelors | 59 | 59 | 2019 | 0·192 | (-0·065, 0·442) | 0·14 | 0·20 |
| | | | | Some college | Bachelors | 34 | 59 | 1125 | 0·167 | (-0·169, 0·471) | 0·33 | 0·40 |
| | Maternal Parity | Yes | 1·94 ± 0·74 | Yes | No | 156 | 68 | 5460 | 0·036 | (-0·174, 0·252) | 0·73 | |
| | | No | 1·86 ± 0·75 | | | | | | | | | |
| | Feeding Type | Exclusive BF | 1·74 ± 0·67 | Combination | Exclusive BF | 41 | 133 | 3135·5 | 0·195 | (-0·063, 0·427) | 0·15 | 0·15 |
| | Feeding Type | Combination | 1·94 ± 0·69 | Combination | Formula | 41 | 49 | 609 | -0·525 | (-0·831, -0·222) | 0·001 | 0·002 |
| | Feeding Type | Formula | 2·39 ± 0·78 | Exclusive BF | Formula | 133 | 49 | 1428 | -0·706 | (-0·917, -0·504) | <0·0001 | <0·0001 |
| | Exposed to Breastmilk | Yes | 1·78 ± 0·68 | No | Yes | 49 | 175 | 6530 | 0·670 | (0·463, 0·882) | <0·0001 | |
| | | No | 2·39 ± 0·78 | | | | | | | | | |
| | Pets in Household | Yes | 1·88 ± 0·70 | No | Yes | 96 | 117 | 6271·5 | 0·155 | (-0·053, 0·346) | 0·14 | 0·14 |
| | | No | 2·00 ± 0·76 | | | | | | | | | |
| M6 | | | | | | | | | | | | |
| | Gestational Age | | 39·69 ± 1·17 | | | | | 1309915·98 | -0·013 | | 0·86 | |
| | Maternal Pre-Pregnancy BMI | | 28·04 ± 7·30 | | | | | 881917·8681 | 0·178 | | 0·0153 | |
| | Weight at Birth (Z-score) | | 0·01 ± 0·86 | | | | | 982731·8327 | 0·084 | | 0·26 | |
| | Breastfeeding Duration at M6 (%) | | 69·52 ± 40·64 | | | | | 1811766·7 | -0·422 | | <0·0001 | |
| | Infant Sex | Female | 2·58 ± 0·69 | Male | Female | 107 | 91 | 4021 | -0·168 | (-0·342, -0·01) | 0·035 | |
| | | Male | 2·44 ± 0·63 | | | | | | | | | |

| Timepoint | Variable | Group | mean±sd | group1 | group2 | n1 | n2 | statistic | estimate | 95%CI | p | p·adj[1] |
|---|---|---|---|---|---|---|---|---|---|---|---|---|
| | Infant Ethnicity | Non-Hispanic or Non-Latino | 2·51 ± 0·66 | Non-Hispanic or Non-Latino | Hispanic or Latino | 180 | 18 | 1620 | 0·000 | (-0·287, 0·292) | 1 | |
| | | Hispanic or Latino | 2·47 ± 0·73 | | | | | | | | | |
| | Infant Ethnicity-Race | White | 2·42 ± 0·62 | Non-White | White | 68 | 130 | 5875 | 0·308 | (0·148, 0·463) | 0·00015 | |
| | | Non-White | 2·67 ± 0·71 | | | | | | | | | |
| | Antibiotics | No | 2·51 ± 0·66 | No | Yes | 189 | 9 | 812 | -0·050 | (-0·506, 0·46) | 0·82 | |
| | | Yes | 2·56 ± 0·71 | | | | | | | | | |
| | Mode of Delivery | Vaginal Delivery | 2·51 ± 0·69 | Vaginal Delivery | C-Section | 152 | 46 | 3684 | 0·051 | (-0·134, 0·241) | 0·58 | |
| | | C-Section | 2·51 ± 0·55 | | | | | | | | | |
| | Enrolled in Medicaid during Pregnancy | No | 2·33 ± 0·64 | No | Yes | 113 | 85 | 2738 | -0·408 | (-0·559, -0·257) | <0·0001 | |
| | | Yes | 2·74 ± 0·62 | | | | | | | | | |
| | Maternal Ethnicity-Race | White | 2·43 ± 0·63 | Non-White | White | 69 | 129 | 5699 | 0·260 | (0·099, 0·423) | 0·0012 | |
| | | Non-White | 2·65 ± 0·70 | | | | | | | | | |
| | Maternal Education | Post graduate degree | 2·27 ± 0·51 | Post graduate degree | <HS or HS | 55 | 62 | 779 | -0·539 | (-0·726, -0·345) | <0·0001 | <0·0001 |
| | | <HS or HS | 2·77 ± 0·61 | Post graduate degree | Some college | 55 | 27 | 383 | -0·500 | (-0·687, -0·255) | 0·000397 | 0·001 |
| | | Some college | 2·61 ± 0·82 | Post graduate degree | Bachelors | 55 | 54 | 1190 | -0·201 | (-0·406, 0·021) | 0·074 | 0·089 |
| | | Bachelors | 2·40 ± 0·67 | <HS or HS | Some college | 62 | 27 | 911·5 | 0·068 | (-0·158, 0·287) | 0·51 | 0·51 |
| | | | | <HS or HS | Bachelors | 62 | 54 | 2308·5 | 0·341 | (0·147, 0·537) | 0·00045 | 0·001 |
| | | | | Some college | Bachelors | 27 | 54 | 960·5 | 0·239 | (0·038, 0·518) | 0·021 | 0·031 |
| | Maternal Parity | No | 2·57 ± 0·57 | Yes | No | 137 | 61 | 3909 | -0·064 | (-0·226, 0·115) | 0·47 | |
| | | Yes | 2·48 ± 0·70 | | | | | | | | | |

| Timepoint | Variable | Group | mean±sd | group1 | group2 | n1 | n2 | statistic | estimate | 95%CI | p | p·adj[1] |
|---|---|---|---|---|---|---|---|---|---|---|---|---|
| | Feeding Type | Exclusive BF | 2·24 ± 0·67 | Combination | Exclusive BF | 35 | 86 | 2007 | 0·321 | (0·101, 0·546) | 0·004 | 0·006 |
| | Feeding Type | Formula | 2·77 ± 0·55 | Combination | Formula | 35 | 76 | 1007 | -0·197 | (-0·382, -0·011) | 0·041 | 0·041 |
| | Feeding Type | Combination | 2·58 ± 0·62 | Exclusive BF | Formula | 86 | 76 | 1609 | -0·512 | (-0·693, -0·328) | <0·0001 | <0·0001 |
| | Exposed to Breastmilk | Yes | 2·34 ± 0·67 | No | Yes | 76 | 121 | 6580 | 0·403 | (0·247, 0·561) | <0·0001 | <0·0001 |
| | | No | 2·77 ± 0·55 | | | | | | | | | |
| | Introduced to Solid Foods | Yes | 2·51 ± 0·67 | Yes | No | 171 | 18 | 1977 | 0·290 | (0·002, 0·582) | 0·048 | |
| | | No | 2·25 ± 0·65 | | | | | | | | | |
| M12 | | | | | | | | | | | | |
| | Gestational Age | | 39·63 ± 1·17 | | | | | 1145426 | -0·121 | | 0·10 | |
| | Maternal Pre-Pregnancy BMI | | 28·24 ± 7·28 | | | | | 836284·5726 | 0·047 | | 0·53 | |
| | Weight at Birth (Z-score) | | -0·00 ± 0·83 | | | | | 839812 | 0·043 | | 0·57 | |
| | Breastfeedinng Duration (weeks) | | 34·43 ± 22·45 | | | | | 1141706·882 | -0·136 | | 0·066 | |
| | Infant Sex | Female | 3·00 ± 0·56 | Male | Female | 96 | 87 | 3125 | -0·218 | (-0·357, -0·071) | 0·0033 | |
| | | Male | 2·78 ± 0·59 | | | | | | | | | |
| | Infant Ethnicity | Non-Hispanic or Non-Latino | 2·88 ± 0·58 | Non-Hispanic or Non-Latino | Hispanic or Latino | 170 | 13 | 1094 | -0·011 | (-0·317, 0·314) | 0·96 | |
| | | Hispanic or Latino | 2·85 ± 0·70 | | | | | | | | | |
| | Infant Ethnicity-Race | Non-White | 3·00 ± 0·62 | Non-White | White | 58 | 125 | 4446 | 0·196 | (0·038, 0·349) | 0·014 | |
| | | White | 2·83 ± 0·56 | | | | | | | | | |
| | Antibiotics | No | 2·88 ± 0·58 | No | Yes | 167 | 16 | 1285 | -0·033 | (-0·294, 0·261) | 0·80 | |
| | | Yes | 2·87 ± 0·64 | | | | | | | | | |

| Timepoint | Variable | Group | mean±sd | group1 | group2 | n1 | n2 | statistic | estimate | 95%CI | p | p·adj[1] |
|---|---|---|---|---|---|---|---|---|---|---|---|---|
| | Mode of Delivery | C-Section | 2·98 ± 0·43 | Vaginal Delivery | C-Section | 136 | 47 | 2963 | -0·062 | (-0·233, 0·093) | 0·46 | |
| | | Vaginal Delivery | 2·85 ± 0·63 | | | | | | | | | |
| | Enrolled in Medicaid during Pregnancy | No | 2·82 ± 0·58 | No | Yes | 110 | 73 | 3349 | -0·150 | (-0·298, 0·005) | 0·058 | |
| | | Yes | 2·97 ± 0·58 | | | | | | | | | |
| | Maternal Ethnicity-Race | White | 2·83 ± 0·56 | Non-White | White | 60 | 123 | 4463 | 0·184 | (0·026, 0·336) | 0·022 | |
| | | Non-White | 2·99 ± 0·63 | | | | | | | | | |
| | Maternal Education | Post graduate degree | 2·80 ± 0·56 | Post graduate degree | <HS or HS | 54 | 49 | 1056 | -0·186 | (-0·374, 0·017) | 0·078 | 0·18 |
| | | Bachelors | 2·84 ± 0·51 | Post graduate degree | Some college | 54 | 25 | 526 | -0·189 | (-0·427, 0·045) | 0·12 | 0·18 |
| | | <HS or HS | 2·94 ± 0·71 | Post graduate degree | Bachelors | 54 | 55 | 1478 | -0·005 | (-0·205, 0·176) | 0·97 | 0·97 |
| | | Some college | 3·03 ± 0·50 | <HS or HS | Some college | 49 | 25 | 607 | -0·007 | (-0·263, 0·236) | 0·95 | 0·97 |
| | | | | <HS or HS | Bachelors | 49 | 55 | 1620 | 0·170 | (-0·019, 0·369) | 0·076 | 0·18 |
| | | | | Some college | Bachelors | 25 | 55 | 839 | 0·179 | (-0·05, 0·425) | 0·12 | 0·18 |
| | Maternal Parity | No | 2·77 ± 0·63 | Yes | No | 128 | 55 | 4093 | 0·151 | (-0·021, 0·326) | 0·081 | |
| | | Yes | 2·93 ± 0·56 | | | | | | | | | |
| | Feeding Type | Still BF | 2·77 ± 0·57 | Still BF | Stopped BF | 83 | 99 | 3147 | -0·210 | (-0·349, -0·06) | 0·007 | |
| | | Stopped BF | 2·98 ± 0·59 | | | | | | | | | |

Abbreviations: Body mass index (BMI); Standard deviation (sd); Number (N); High School or less than high-school (<HS or HS); Cesarian section (C-Section); Breastfeeding (BF); Month one visit (M1); Month six visit (M6); Month twelve visit (M12); Adjusted P-value (p.adj)

[1]Benjamini-Hochberg (BH) corrected

Supplemental Table 3 Mean relative abundance of aggregated taxa over the first year of life.

| Timepoint | Phylum | Genus | Genus·Agg | Mean Relative Abundance |
|---|---|---|---|---|
| Birth | Actinobacteriota | *Actinomyces* | Other | 0·0925 |
| Birth | Firmicutes | *Anaerococcus* | Anaerococcus | 0·4774 |
| Birth | Firmicutes | *Anaerostipes* | Other | 0·0191 |
| Birth | Actinobacteriota | *Atopobium* | Other | 0·0064 |
| Birth | Bacteroidota | *Bacteroides* | Bacteroides | 2·9896 |
| Birth | Actinobacteriota | *Bifidobacterium* | Bifidobacterium | 3·2724 |
| Birth | Firmicutes | *Blautia* | Other | 0·0107 |
| Birth | Firmicutes | *Clostridioides* | Other | 0·0001 |
| Birth | Firmicutes | *Clostridium_innocuum_group* | Other | 0·0033 |
| Birth | Firmicutes | *Clostridium_sensu_stricto_1* | Other | 1·8363 |
| Birth | Actinobacteriota | *Collinsella* | Collinsella | 0·0000 |
| Birth | Actinobacteriota | *Corynebacterium* | Corynebacterium | 9·1817 |
| Birth | Firmicutes | *Dialister* | Other | 0·0029 |
| Birth | Actinobacteriota | *Eggerthella* | Other | 0·0010 |
| Birth | Firmicutes | *Enterococcus* | Enterococcus | 11·2188 |
| Birth | Firmicutes | *Erysipelatoclostridium* | Other | 0·1233 |
| Birth | Proteobacteria | *Escherichia-Shigella* | Escherichia-Shigella | 22·7787 |
| Birth | Firmicutes | *Ezakiella* | Other | 0·1561 |
| Birth | Firmicutes | *Faecalibacterium* | Other | 0·2354 |
| Birth | Firmicutes | *Fenollaria* | Other | 0·0089 |
| Birth | Firmicutes | *Finegoldia* | Finegoldia | 0·5754 |
| Birth | Firmicutes | *Flavonifractor* | Other | 0·0000 |
| Birth | Proteobacteria | *Haemophilus* | Haemophilus | 0·0125 |
| Birth | Firmicutes | *Incertae_Sedis* | Other | 0·0107 |
| Birth | Firmicutes | *Intestinibacter* | Other | 0·0008 |
| Birth | Proteobacteria | *Klebsiella* | Klebsiella | 0·6462 |
| Birth | Firmicutes | *Lachnoclostridium* | Other | 0·0843 |
| Birth | Firmicutes | *Lactobacillus* | Other | 1·7194 |
| Birth | Actinobacteriota | *Lawsonella* | Lawsonella | 0·0153 |
| Birth | Firmicutes | *Megasphaera* | Other | 0·0497 |
| Birth | Firmicutes | *Mogibacterium* | Other | 0·0000 |
| Birth | Firmicutes | *Murdochiella* | Other | 0·0421 |
| Birth | Firmicutes | *Negativicoccus* | Other | 0·0043 |
| Birth | Bacteroidota | *Parabacteroides* | Parabacteroides | 0·5067 |
| Birth | Firmicutes | *Peptococcus* | Other | 0·0036 |
| Birth | Firmicutes | *Peptoniphilus* | Peptoniphilus | 0·6221 |
| Birth | Firmicutes | *Peptostreptococcus* | Other | 0·1571 |
| Birth | Bacteroidota | *Porphyromonas* | Porphyromonas | 0·0008 |

| Timepoint | Phylum | Genus | Genus·Agg | Mean Relative Abundance |
|---|---|---|---|---|
| Birth | Bacteroidota | *Prevotella* | Prevotella | 0·3678 |
| Birth | Firmicutes | *Ruminococcus_gnavus_group* | Other | 0·1372 |
| Birth | Firmicutes | *Ruminococcus_torques_group* | Other | 0·0027 |
| Birth | Firmicutes | *S5-A14a* | Other | 0·0459 |
| Birth | Firmicutes | *Sellimonas* | Other | 0·0281 |
| Birth | Firmicutes | *Staphylococcus* | Staphylococcus | 32·6382 |
| Birth | Firmicutes | *Streptococcus* | Streptococcus | 9·2797 |
| Birth | Proteobacteria | *Sutterella* | Sutterella | 0·0064 |
| Birth | Firmicutes | *Tyzzerella* | Other | 0·0008 |
| Birth | Actinobacteriota | *Varibaculum* | Varibaculum | 0·0514 |
| Birth | Firmicutes | *Veillonella* | Other | 0·5764 |
| M1 | Actinobacteriota | *Actinomyces* | Other | 1·0316 |
| M1 | Firmicutes | *Anaerococcus* | Anaerococcus | 6·4121 |
| M1 | Firmicutes | *Anaerostipes* | Other | 0·0152 |
| M1 | Actinobacteriota | *Atopobium* | Other | 0·2521 |
| M1 | Bacteroidota | *Bacteroides* | Bacteroides | 3·7763 |
| M1 | Actinobacteriota | *Bifidobacterium* | Bifidobacterium | 23·0340 |
| M1 | Firmicutes | *Blautia* | Other | 0·4381 |
| M1 | Firmicutes | *Clostridioides* | Other | 0·0883 |
| M1 | Firmicutes | *Clostridium_innocuum_group* | Other | 0·0815 |
| M1 | Firmicutes | *Clostridium_sensu_stricto_1* | Other | 4·7116 |
| M1 | Actinobacteriota | *Collinsella* | Collinsella | 0·9167 |
| M1 | Actinobacteriota | *Corynebacterium* | Corynebacterium | 0·8280 |
| M1 | Firmicutes | *Dialister* | Other | 0·5329 |
| M1 | Actinobacteriota | *Eggerthella* | Other | 0·1613 |
| M1 | Firmicutes | *Enterococcus* | Enterococcus | 3·4250 |
| M1 | Firmicutes | *Erysipelatoclostridium* | Other | 1·3714 |
| M1 | Proteobacteria | *Escherichia-Shigella* | Escherichia-Shigella | 7·1946 |
| M1 | Firmicutes | *Ezakiella* | Other | 0·2391 |
| M1 | Firmicutes | *Faecalibacterium* | Other | 0·2696 |
| M1 | Firmicutes | *Fenollaria* | Other | 0·2999 |
| M1 | Firmicutes | *Finegoldia* | Finegoldia | 11·5208 |
| M1 | Firmicutes | *Flavonifractor* | Other | 0·0958 |
| M1 | Proteobacteria | *Haemophilus* | Haemophilus | 0·3474 |
| M1 | Firmicutes | *Incertae_Sedis* | Other | 0·0058 |
| M1 | Firmicutes | *Intestinibacter* | Other | 0·1687 |
| M1 | Proteobacteria | *Klebsiella* | Klebsiella | 1·2632 |
| M1 | Firmicutes | *Lachnoclostridium* | Other | 0·3081 |
| M1 | Firmicutes | *Lactobacillus* | Other | 1·1798 |
| M1 | Actinobacteriota | *Lawsonella* | Lawsonella | 1·7206 |

| Timepoint | Phylum | Genus | Genus·Agg | Mean Relative Abundance |
|---|---|---|---|---|
| M1 | Firmicutes | *Megasphaera* | Other | 0·8739 |
| M1 | Firmicutes | *Mogibacterium* | Other | 0·0480 |
| M1 | Firmicutes | *Murdochiella* | Other | 0·2228 |
| M1 | Firmicutes | *Negativicoccus* | Other | 0·4963 |
| M1 | Bacteroidota | *Parabacteroides* | Parabacteroides | 0·3128 |
| M1 | Firmicutes | *Peptococcus* | Other | 0·1230 |
| M1 | Firmicutes | *Peptoniphilus* | Peptoniphilus | 6·9115 |
| M1 | Firmicutes | *Peptostreptococcus* | Other | 0·7633 |
| M1 | Bacteroidota | *Porphyromonas* | Porphyromonas | 0·7371 |
| M1 | Bacteroidota | *Prevotella* | Prevotella | 3·5515 |
| M1 | Firmicutes | *Ruminococcus_gnavus_group* | Other | 1·1591 |
| M1 | Firmicutes | *Ruminococcus_torques_group* | Other | 0·3043 |
| M1 | Firmicutes | *S5-A14a* | Other | 0·0917 |
| M1 | Firmicutes | *Sellimonas* | Other | 0·0965 |
| M1 | Firmicutes | *Staphylococcus* | Staphylococcus | 0·8214 |
| M1 | Firmicutes | *Streptococcus* | Streptococcus | 7·6746 |
| M1 | Proteobacteria | *Sutterella* | Sutterella | 0·0631 |
| M1 | Firmicutes | *Tyzzerella* | Other | 0·0287 |
| M1 | Actinobacteriota | *Varibaculum* | Varibaculum | 1·1766 |
| M1 | Firmicutes | *Veillonella* | Other | 2·8542 |
| M6 | Actinobacteriota | *Actinomyces* | Other | 1·0696 |
| M6 | Firmicutes | *Anaerococcus* | Anaerococcus | 7·5516 |
| M6 | Firmicutes | *Anaerostipes* | Other | 0·4258 |
| M6 | Actinobacteriota | *Atopobium* | Other | 0·9535 |
| M6 | Bacteroidota | *Bacteroides* | Bacteroides | 2·8600 |
| M6 | Actinobacteriota | *Bifidobacterium* | Bifidobacterium | 18·2879 |
| M6 | Firmicutes | *Blautia* | Other | 2·0523 |
| M6 | Firmicutes | *Clostridioides* | Other | 0·2301 |
| M6 | Firmicutes | *Clostridium_innocuum_group* | Other | 0·3350 |
| M6 | Firmicutes | *Clostridium_sensu_stricto_1* | Other | 0·3827 |
| M6 | Actinobacteriota | *Collinsella* | Collinsella | 1·8248 |
| M6 | Actinobacteriota | *Corynebacterium* | Corynebacterium | 0·1769 |
| M6 | Firmicutes | *Dialister* | Other | 1·7754 |
| M6 | Actinobacteriota | *Eggerthella* | Other | 0·4195 |
| M6 | Firmicutes | *Enterococcus* | Enterococcus | 2·1991 |
| M6 | Firmicutes | *Erysipelatoclostridium* | Other | 0·4531 |
| M6 | Proteobacteria | *Escherichia-Shigella* | Escherichia-Shigella | 3·0941 |
| M6 | Firmicutes | *Ezakiella* | Other | 0·3619 |
| M6 | Firmicutes | *Faecalibacterium* | Other | 0·5488 |
| M6 | Firmicutes | *Fenollaria* | Other | 1·7087 |

| Timepoint | Phylum | Genus | Genus·Agg | Mean Relative Abundance |
|---|---|---|---|---|
| M6 | Firmicutes | *Finegoldia* | Finegoldia | 11·6082 |
| M6 | Firmicutes | *Flavonifractor* | Other | 0·1868 |
| M6 | Proteobacteria | *Haemophilus* | Haemophilus | 0·1060 |
| M6 | Firmicutes | *Incertae_Sedis* | Other | 0·0701 |
| M6 | Firmicutes | *Intestinibacter* | Other | 0·5957 |
| M6 | Proteobacteria | *Klebsiella* | Klebsiella | 0·1261 |
| M6 | Firmicutes | *Lachnoclostridium* | Other | 0·6839 |
| M6 | Firmicutes | *Lactobacillus* | Other | 0·5110 |
| M6 | Actinobacteriota | *Lawsonella* | Lawsonella | 1·0580 |
| M6 | Firmicutes | *Megasphaera* | Other | 1·2840 |
| M6 | Firmicutes | *Mogibacterium* | Other | 0·1815 |
| M6 | Firmicutes | *Murdochiella* | Other | 1·0923 |
| M6 | Firmicutes | *Negativicoccus* | Other | 0·9321 |
| M6 | Bacteroidota | *Parabacteroides* | Parabacteroides | 0·2389 |
| M6 | Firmicutes | *Peptococcus* | Other | 0·3277 |
| M6 | Firmicutes | *Peptoniphilus* | Peptoniphilus | 9·2690 |
| M6 | Firmicutes | *Peptostreptococcus* | Other | 1·8001 |
| M6 | Bacteroidota | *Porphyromonas* | Porphyromonas | 1·6353 |
| M6 | Bacteroidota | *Prevotella* | Prevotella | 6·7692 |
| M6 | Firmicutes | *Ruminococcus_gnavus_group* | Other | 2·9091 |
| M6 | Firmicutes | *Ruminococcus_torques_group* | Other | 0·8236 |
| M6 | Firmicutes | *S5-A14a* | Other | 0·6093 |
| M6 | Firmicutes | *Sellimonas* | Other | 0·3503 |
| M6 | Firmicutes | *Staphylococcus* | Staphylococcus | 0·3418 |
| M6 | Firmicutes | *Streptococcus* | Streptococcus | 3·1847 |
| M6 | Proteobacteria | *Sutterella* | Sutterella | 0·1372 |
| M6 | Firmicutes | *Tyzzerella* | Other | 0·8713 |
| M6 | Actinobacteriota | *Varibaculum* | Varibaculum | 1·8528 |
| M6 | Firmicutes | *Veillonella* | Other | 3·7332 |
| M12 | Actinobacteriota | *Actinomyces* | Other | 0·6868 |
| M12 | Firmicutes | *Anaerococcus* | Anaerococcus | 7·8330 |
| M12 | Firmicutes | *Anaerostipes* | Other | 1·4855 |
| M12 | Actinobacteriota | *Atopobium* | Other | 0·8204 |
| M12 | Bacteroidota | *Bacteroides* | Bacteroides | 3·1292 |
| M12 | Actinobacteriota | *Bifidobacterium* | Bifidobacterium | 10·8435 |
| M12 | Firmicutes | *Blautia* | Other | 4·4764 |
| M12 | Firmicutes | *Clostridioides* | Other | 0·1990 |
| M12 | Firmicutes | *Clostridium_innocuum_group* | Other | 0·1881 |
| M12 | Firmicutes | *Clostridium_sensu_stricto_1* | Other | 0·4767 |
| M12 | Actinobacteriota | *Collinsella* | Collinsella | 1·4986 |

| Timepoint | Phylum | Genus | Genus·Agg | Mean Relative Abundance |
|---|---|---|---|---|
| M12 | Actinobacteriota | *Corynebacterium* | Corynebacterium | 0·7820 |
| M12 | Firmicutes | *Dialister* | Other | 2·5395 |
| M12 | Actinobacteriota | *Eggerthella* | Other | 0·3158 |
| M12 | Firmicutes | *Enterococcus* | Enterococcus | 2·0293 |
| M12 | Firmicutes | *Erysipelatoclostridium* | Other | 0·5514 |
| M12 | Proteobacteria | *Escherichia-Shigella* | Escherichia-Shigella | 1·1727 |
| M12 | Firmicutes | *Ezakiella* | Other | 1·4196 |
| M12 | Firmicutes | *Faecalibacterium* | Other | 4·1172 |
| M12 | Firmicutes | *Fenollaria* | Other | 2·3188 |
| M12 | Firmicutes | *Finegoldia* | Finegoldia | 11·1001 |
| M12 | Firmicutes | *Flavonifractor* | Other | 0·1866 |
| M12 | Proteobacteria | *Haemophilus* | Haemophilus | 0·0930 |
| M12 | Firmicutes | *Incertae_Sedis* | Other | 0·1119 |
| M12 | Firmicutes | *Intestinibacter* | Other | 0·4981 |
| M12 | Proteobacteria | *Klebsiella* | Klebsiella | 0·2502 |
| M12 | Firmicutes | *Lachnoclostridium* | Other | 0·6436 |
| M12 | Firmicutes | *Lactobacillus* | Other | 0·2162 |
| M12 | Actinobacteriota | *Lawsonella* | Lawsonella | 0·7101 |
| M12 | Firmicutes | *Megasphaera* | Other | 0·6544 |
| M12 | Firmicutes | *Mogibacterium* | Other | 0·5780 |
| M12 | Firmicutes | *Murdochiella* | Other | 1·1055 |
| M12 | Firmicutes | *Negativicoccus* | Other | 0·6762 |
| M12 | Bacteroidota | *Parabacteroides* | Parabacteroides | 0·2995 |
| M12 | Firmicutes | *Peptococcus* | Other | 0·2280 |
| M12 | Firmicutes | *Peptoniphilus* | Peptoniphilus | 9·4489 |
| M12 | Firmicutes | *Peptostreptococcus* | Other | 2·3797 |
| M12 | Bacteroidota | *Porphyromonas* | Porphyromonas | 2·6424 |
| M12 | Bacteroidota | *Prevotella* | Prevotella | 7·8362 |
| M12 | Firmicutes | *Ruminococcus_gnavus_group* | Other | 2·3750 |
| M12 | Firmicutes | *Ruminococcus_torques_group* | Other | 0·8013 |
| M12 | Firmicutes | *S5-A14a* | Other | 0·8989 |
| M12 | Firmicutes | *Sellimonas* | Other | 0·3458 |
| M12 | Firmicutes | *Staphylococcus* | Staphylococcus | 1·1182 |
| M12 | Firmicutes | *Streptococcus* | Streptococcus | 2·1897 |
| M12 | Proteobacteria | *Sutterella* | Sutterella | 0·3970 |
| M12 | Firmicutes | *Tyzzerella* | Other | 0·8561 |
| M12 | Actinobacteriota | *Varibaculum* | Varibaculum | 2·5297 |
| M12 | Firmicutes | *Veillonella* | Other | 1·9459 |

Abbreviations: Month 1 visit (M1); Month 6 visit (M6); Month 12 visit (M12); Aggregate (Agg)

Supplemental Table 4 Demographics of infants part of full Rotavirus-IgA cohort and of Sub-cohort

| Characteristic | Rotavirus-IgA cohort N = 65[1] | Sub-cohort† N = 47[1] | p-value[2] |
|---|---|---|---|
| **Infant Sex** | | | 0·87 |
| Male | 39/65 (60%) | 29/47 (62%) | |
| Female | 26/65 (40%) | 18/47 (38%) | |
| **Infant Ethnicity-Race** | | | 0.91 |
| Non-White | 27/65 (42%) | 19/47 (40%) | |
| White | 38/65 (58%) | 28/47 (60%) | |
| **Delivery Mode** | | | 0·74 |
| Vaginal Delivery | 48/65 (74%) | 36/47 (77%) | |
| C-Section | 17/65 (26%) | 11/47 (23%) | |
| **Gestational Age (weeks)** | 39·712 ± 1·152 | 39·562 ± 1·131 | 0·53 |
| **Birth Weight (Z - score)** | -0·027 ± 0·861 | -0·082 ± 0·885 | 0·87 |
| **Antibiotics (previous to M1 visit)** | | | >0·99 |
| No | 61/62 (98%) | 47/47 (100%) | |
| Yes | 1/62 (1·6%) | 0/47 (0%) | |
| Unknown | 3 | 0 | |
| **Feeding Type (Month 1)** | | | 0·87 |
| Combination | 12/62 (19%) | 10/47 (21%) | |
| Exclusive BF | 34/62 (55%) | 27/47 (57%) | |
| Formula | 16/62 (26%) | 10/47 (21%) | |
| Unknown | 3 | 0 | |
| **Age at First Vaccine Dose (weeks)** | 9·800 ± 1·743 | 9·426 ± 0·617 | 0·53 |
| **Product of First Dose** | | | >0·99 |
| Rotarix | 1/65 (1·5%) | 0/47 (0%) | |
| RotaTeq | 64/65 (98%) | 47/47 (100%) | |
| **Full Vaccination Status By M6 Visit** | | | 0·71 |
| Full schedule | 30/65 (46%) | 20/47 (43%) | |
| Partial schedule | 35/65 (54%) | 27/47 (57%) | |
| **Weeks Between Dose & Month One Rectal Swab Collection[3]** | 4·113 ± 2·146 | 3·916 ± 1·882 | 0.91 |

Abbreviations: Breastfeeding (BF); Cesarean Section (C-Section); Month six (M6)

[1]n/N (%); Mean ± SD

[2]Pearson's Chi-squared test; Fisher's exact test; Wilcoxon rank sum test

[3]Positive values represent dose given after sample collection

† Sub-cohort: Subset of infants within the full cohort who had both a M6 Rotavirus-IgA measurement and a M1 microbiome sample·

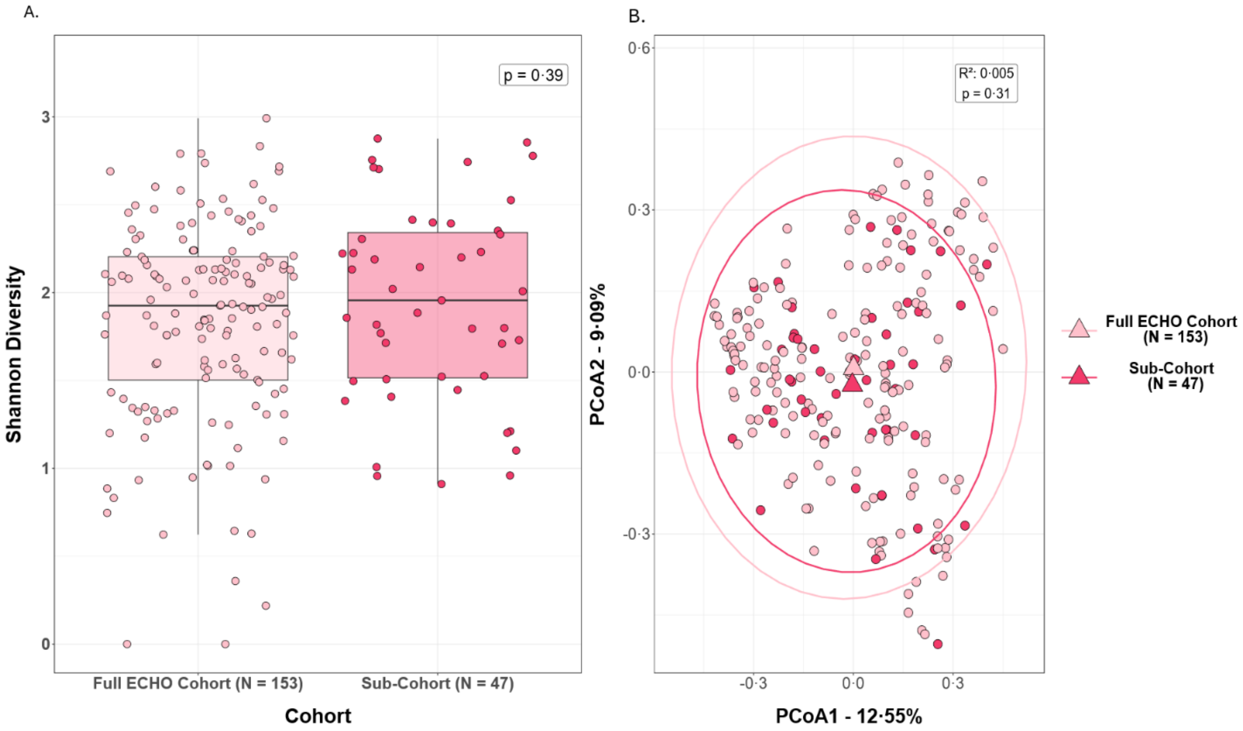

**Supplemental Figure 1 Differences in alpha and beta diversity at M1 between full ECHO-Cohort and Sub-Cohort**

A) Shannon diversity index at M1 visit for the Full ECHO cohort and the Sub-cohort· Each boxplot represents the Shannon diversity for the respective cohort· Differences tested through Wilcoxon test· B) Beta diversity at M1 visit was determined using Bray-Curtis dissimilarity metric· Centroids of each group are represented by a triangle· Differences tested through PERMANOVA.
